# Systematic review and meta-analysis of the association between Epstein-Barr virus, Multiple Sclerosis, and other risk factors

**DOI:** 10.1101/19007450

**Authors:** Benjamin M Jacobs, Gavin Giovannoni, Jack Cuzick, Ruth Dobson

## Abstract

**Background:** EBV infection is thought to play a central role in the development of Multiple Sclerosis (MS). If causal, it represents a target for interventions to reduce MS risk.

**Objective:** To examine the evidence for interaction between EBV and other risk factors, and explore mechanisms via which EBV infection may influence MS risk.

**Methods:** Pubmed was searched using the terms “multiple sclerosis” AND “Epstein Barr virus”, “multiple sclerosis” AND EBV, “clinically isolated syndrome” AND “Epstein Barr virus” and “clinically isolated syndrome” AND EBV. All abstracts were reviewed for possible inclusion.

**Results:** 262 full-text papers were reviewed. There was evidence of interaction on the additive scale between anti-EBV antibody titre and HLA genotype (AP 0.48, p<1×10^−4^; RERI 3.84, p<5×10^−3^; S 1.68, p=0.06). Previous IM was associated with increased OR of MS in HLA-DRB1*1501 positive but not HLA-DRB1*1501 negative persons. Smoking was associated with a greater risk of MS in those with high anti-EBV antibodies (OR 2.76) but not low anti-EBV antibodies (OR 1.16). No interaction between EBV and risk factors was found on a multiplicative scale.

**Conclusions:** EBV appears to interact with at least some established MS risk factors. The mechanism via which EBV influences MS risk remains unknown.

## Introduction

Multiple sclerosis (MS) is thought to arise as the result of acquired environmental risk in a genetically susceptible population^1–4^. Environmental risk factors for MS include Epstein-Barr Virus (EBV) infection, smoking, obesity during adolescence, and low serum vitamin D^2^. Understanding how environmental risk factors interact with each other and with genotype is crucial to developing targeted preventative strategies.

We set out to update and extend our understanding of the interaction between EBV and other MS risk factors. To our knowledge, there has been no previous attempt to integrate all data related to how EBV interacts with other MS risk factors. One meta-analysis has examined the potential interaction between EBV serostatus and HLA in MS; other previous meta-analyses have not studied risk factor interaction^5–8^.

Nested case-control studies using large health repositories^9,10^ have made a major contribution to epidemiological evidence supporting a causal relationship between EBV and MS. However, the high rate of EBV seropositivity in the general population argues against EBV seropositivity alone being a sufficient factor for causing MS^7^. The prevalence of MS in EBV-negative individuals is virtually zero when highly sensitive techniques are used to assess EBV serostatus^11,12^. Symptomatic EBV^5–7,13–15^ infection (Infectious Mononucleosis; IM) confers a greater risk of MS than asymptomatic EBV carriage.

Population-based epidemiological studies indicate that EBV infection and other environmental risk factors may interact with genotype in the pathogenesis of MS ^16^. To our knowledge there have been no previous attempts to systematically pool these estimates. In this systematic review and meta-analysis, we examine all the available evidence for EBV interaction with other MS risk factors (both in terms of EBV serostatus and IM) using both multiplicative and additive models for interaction. We also examine the reported relationship between EBV and MS, and pool evidence around the relationship between active EBV turnover (as measured by PCR) and MS. Finally, we provide a narrative systematic review of the literature around MS and EBV.

## Methods

### Search strategy

Pubmed was searched using the terms “multiple sclerosis” AND “Epstein Barr virus”, “multiple sclerosis” AND EBV, “clinically isolated syndrome” AND “Epstein Barr virus” and “clinically isolated syndrome” AND EBV. Search dates were 1950-present. The most recent search was performed on 22nd December 2018.

All abstracts were reviewed for possible inclusion. Studies for use in the meta-analysis were screened according to the following criteria: containing both MS and control group, and using either standard techniques to establish EBV serostatus, history of IM, or PCR. Where these criteria were met, the full text was retrieved.

Following this, relevant studies were reviewed and data extracted. Where full text was not available, the authors were contacted to provide the article. Where it was judged unclear as to whether data within selected papers met the inclusion criteria (details of inclusion criteria for each analysis are given in the results section), a second co-author independently reviewed the paper, and a consensus decision was reached. The quality of data were assessed by recording the reported security of MS diagnosis (no clear criteria and/or self-reported vs. explicit criteria used for diagnosis, the gold standard), and technique for assessing EBV (ELISA vs. immunofluorescence, the gold standard). All references of retrieved review and/or meta-analyses were reviewed for additional articles not captured during the original search.

Technical differences between study design may introduce bias and limit the validity of pooled effect estimates. Such differences included: differences in clinical criteria for MS diagnosis, differences in method of IM diagnosis (clinical, recall questionnaire, serological), differences in laboratory techniques (e.g. immunoflourescence vs ELISA), differences in HLA genotyping (molecular typing vs SNP imputation), and difference in the quantification of smoking exposure (cotinine vs questionnaire). To overcome these difficulties, we performed subgroup analyses where appropriate to stratify by these potential sources of heterogeneity.

All included full text papers were assigned to analyses covered by this review - EBV interaction with other MS risk factors, serology and MS risk, infectious mononucleosis and MS risk, EBV DNA detection and MS, papers covering possible mechanisms of EBV contribution to MS risk, and papers examining the relationship between immune response to EBV and MS-related clinical or MRI outcomes. A single paper could be assigned to any number of analyses, and each analysis/review was performed independently of all others.

### Statistical methods

Meta-analyses were conducted in R v3.6.0 using the ‘meta’ package based on reported data. Odds ratios (ORs) were calculated using a Mantel-Haenszel random effects model with a continuity correction. Bias was quantified using the efficient score (a linear regression of funnel plot asymmetry)^17^. For interaction studies, odds ratios were pooled using the inverse variance method. Where data were available, unadjusted odds ratios were calculated.

For interaction studies, the highest and lowest exposure groups were used - e.g. where EBNA titres were divided into quartiles, we took the lowest and the highest groups as ‘EBNA lo’ and ‘EBNA hi’ respectively. Interaction was assessed by calculating 4 measures of interaction: where the numbers of cases and controls in each risk factor group were presented, the Attributable Proportion due to interaction (AP), the relative excess risk due to interaction (RERI), the Synergy index (S), and multiplicative interaction^18,19^ were calculated. For two risk factors of interest, e.g. smoking and HLA status, if OR_11_ indicates the Odds Ratio for MS in individuals exposed to both risk factors, OR_10_ the OR for HLA+ non-smokers and OR_01_ that for HLA-smokers:

RERI = OR_11_ – OR_10_ – OR_01_ + 1

S = OR_11_ – 1 / ((OR_10_ – 1) – (OR_01_ – 1))

AP = (OR_11_ – OR_10_ – OR_01_ + 1)/ OR_11_

In the absence of interaction, RERI and AP will be 0, and S will be 1. Measures of departure from additivity (AP, RERI, and S) were calculated using the indicator variable method described previously^19^ in R (code available on request). Multiplicative interaction was calculated by performing logistic regression with an interaction term. If OR represents the Odds Ratio for MS, x_1_ one risk factor, x_2_ the second risk factor, and x_1_x_2_ the product (interaction) term, then:

Ln(OR) = b_0_ + b_1_x_1_ + b_2_x_2_ + b_3_x_1_x_2_

The exponent of the interaction term coefficient b_3_ represents the multiplicative interaction between the two risk factors. For these analyses, the regression model did not adjust for variables other than the two risk factors in question. Standard errors were calculated for measures of additive interaction using the delta method^19^. Standard errors for the multiplicative interaction were calculated from the output of the logistic regression model. Meta-analysis of interaction terms was performed using the inverse variance method with a random effects model.

### Data availability statement

This work was performed using published data. All data sources are listed in the references and supplementary references. All R code used for the analysis are available on request.

## Results

A total of 632 references were retrieved using the search terms “multiple sclerosis” AND “Epstein Barr virus”, and “multiple sclerosis” AND EBV. “Multiple sclerosis” AND EBV, “clinically isolated syndrome” retrieved 22 references, all of which had been captured in the previous search. “Clinically isolated syndrome” AND EBV retrieved a further 17 references, again all of which had been previously captured. Review of all references of meta-analyses and systematic reviews provided 6 unique new results. 370 results were discarded following review of abstracts for reasons including pre-selecting EBV positive patients only, having no control group, validation studies of new methods for EBV serology. 262 full text papers were reviewed and included as summarized in Fig. 1.

### EBV interaction with HLA-DRB1*1501 in MS

10 papers^20–28^ were included for this analysis. All but one paper presented HLA-DRB1*1501 homo- and heterozygotes pooled into a single group (“HLA positive”), and so this grouping was used in the analysis. Where EBNA titres were divided into quartiles, we took the highest and lower quartiles to represent ‘high’ and ‘low’ titres respectively. One paper^26^ was excluded due to overlapping participants with another paper^22^.

The odds ratio (OR) of MS in individuals with high anti-EBV antibody titres is increased in HLA-DRB1*1501 positive (OR 7.17, 95% CI 3.92 - 13.14) compared to HLA-DRB1*1501 negative individuals (OR 2.92, 95% CI 1.86 - 4.59, Fig. 2, Table 1). Studies differed in their method of HLA genotyping. Restricting the analysis to studies using tagging SNPs (rs3135005 or rs9271366) did not significantly alter the results (Fig. S1). Restricting the analysis to studies using PCR-based methods yielded a similar result (Fig. S1).

Individual-level data were available for five studies. We estimated the degree of interaction between HLA status and EBNA titre by calculating the AP, Synergy Index, RERI, and the degree of multiplicative interaction as described above. There was evidence of significant interaction between EBNA titre and HLA genotype on the additive scale in terms of the AP and RERI (AP 0.48, p<1×10^−4^; RERI 3.84, p<5×10^−3^; S 1.68, p=0.06). There was no evidence of interaction on the multiplicative scale (/3 1.27, p=0.74) (Fig. 2, Table 1). Subgroup analyses based on method of HLA genotyping are presented in table S1.

Four papers examined the potential interaction between previous infectious mononucleosis and HLA-DRB1*1501 status and MS^20,22,28,29^. Again, homo- and heterozygote status was pooled into “HLA positive”. A history of IM is associated with increased OR of MS in HLA-DRB1*1501 positive individuals (OR 5.11 95% CI 2.00-13.03; p<1×10^−3^) but not in HLA-DRB1*1501 negative individuals (OR 1.22 95% CI 0.33-4.48; p=0.77, Fig. 3, Table 2).

Three studies had individual-level data available. There was no significant interaction on the additive or multiplicative scales between HLA status and IM in the meta-analysis of 3 studies with individual-level data available (figure 3, Table 2). Subgroup analysis by method of HLA genotyping did not significantly alter the results (Fig. S2).

### EBV interaction with smoking in MS

5 papers studied the potential interaction between smoking status and anti-EBV antibody titre^25,26,28,30,31^. Three studies stratified smoke exposure as ever vs never smokers, one study used second-hand smoke exposure as a variable, and one study distinguished active from inactive smoking using serum cotinine levels. Smoking is associated with a greater risk of MS in those with high anti-EBV antibodies (OR 2.76 95%CI 2.13-3.59; p<1×10^−5^) but not in those with low anti-EBV antibodies (OR 1.16 95%CI 0.95-1.42; p=0.15). There was no significant interaction on the multiplicative or additive scales in the meta-analysis of the four eligible studies (Fig. 4, Table 3). Exclusion of either the study using second-hand smoke as the exposure or using serum cotinine as a proxy for smoking did not significantly affect the results (Table S2).

### EBV interaction with vitamin D in MS risk

Only 2 studies presented data on both EBV and vitamin D in MS^32,33^. One of these studies looked at vitamin D levels in people with established MS^32^, and the other in samples taken both prior to and following MS onset, with multiple, variable sampling points per participant^33^. One study applied a correction to vitamin D levels for month of sampling^33^, the other did not^32^. In addition, one study using a single EBNA epitope^33^, whereas the other looked at specific ENA-1 domains^32^. Neither study demonstrated any interaction between vitamin D level and anti-EBNA titre, however for the reasons above they were not pooled.

### EBV interaction with obesity in MS risk

Only one study examined the potential interaction between EBV and obesity in risk of MS^34^. This study demonstrated a striking potential interaction on an additive scale with an attributable proportion due to interaction of 0.8 (95%CI 0.6-1.0) in the incident study, and in the prevalent study an attributable proportion due to interaction of 0.7 (95%CI 0.5-1.0)^34^.

### EBV seropositivity and MS

56 papers were included in the final analysis for this analysis (supplementary references). Inclusion criteria were MS and control group, no pre-selection of groups based on EBV serostatus and history of IM, EBV serology measured using clearly defined methods. Reasons for exclusion included not having a control group and pre-selecting EBV positive patients.

Studies were separated into those examining adult vs. paediatric MS populations given the reported differences in seroprevalence between the two groups. Following an assessment of data quality, validatory analyses were performed limiting studies to those deemed to be of high quality. Seropositivity for EBV was calculated by pooling results from studies which reported seropositivity to either EBNA, VCA, or both. Where both were reported, the EBNA data were used. Studies using different EBNA1 and EBNA2 epitopes were pooled for all analysis. EBV seropositivity was significantly more common among people with MS (adults and children) than controls (n=15554, 56 studies, OR 3.62, 95% CI 2.83-4.83, Fig. 5). There was evidence of significant heterogeneity (Q=168.37, p<1×10^−4^) and publication bias (p<0.05).

Overall, 6479/7395 people with MS were EBV seropositive (87.6%) compared with 6205/8159 EBV seropositive control subjects (76.1%). EBV seropositivity was more prevalent among adults with MS compared to controls (n=13768, 46 studies, OR 3.27, 95% CI 2.45-4.35, p<1×10^−4^). There was substantial heterogeneity between studies (Q=134.5 p<×10^−4^) and evidence of publication bias (p=0.0196), with studies demonstrating a relationship between EBV infection and MS more likely to be published. Overall, 5805/6606 adults with MS were EBV seropositive (87.9%) compared with 5702/7162 adult control subjects (79.6%). EBV seropositivity was more common among children with MS or CIS than controls (n=1786, 10 studies, OR 4.93, 95% CI 3.83-6.36, p<1×10^−4^). There was no evidence of heterogeneity (Q=5.76, p=0.76) and no evidence of publication bias (p=0.622). Overall, 674/789 children with MS were EBV seropositive (85.4%) compared with 485/997 control subjects (48.6%).

IgG reactivity to the Viral Capsid Antigen (VCA) was more prevalent among adults with MS (n=7688, 23 studies, OR 3.18, 95% CI 1.88-5.38, p<1×10^−4^, data not shown). There was substantial heterogeneity between studies (Q=73.27, p<1×10^−4^) and no evidence of publication bias (p=0.062). Overall, VCA seropositivity was present in 3414/3698 (92.3%) of adults with MS compared with 3595/3990 controls (90.1%). Reactivity to the EBNA antigen was again more prevalent among people with MS compared to controls (n=9537, 33 studies, OR 3.39, 95% CI 2.53-4.56, p<1×10^−4^, data not shown). There was substantial heterogeneity between studies (Q=71.5, p<1×10^−4^) with evidence of publication bias in these studies (p<0.01). Overall, EBNA seropositivity was present in 4260/4805 (88.7%) of adults with MS compared with 3660/4732 (77.3%) controls.

The increased seroprevalence of EBV infection in people with MS/CIS remained significant when restricting included studies to those using the more sensitive technique of immunofluorescence (rather than enzyme-linked immunosorbent assay) to detect EBV antibodies (OR 4.61, 95% CI 1.86-11.40). Similarly, when restricting included studies to those which used explicit diagnostic criteria to define MS, this effect remained significant (OR 3.44, 95% CI 2.56-4.61)

### Infectious mononucleosis and MS

32 full text papers were reviewed for this analysis, of which 19 met the inclusion criteria (supplementary references). Inclusion criteria were MS and control group, clearly stated methods for obtaining a previous history of IM, and no selection on the basis of reported history of IM.

Previous Infectious Mononucleosis (IM) was more common in people with MS (n=107243, 19 studies, OR 2.00, 95% CI 1.79-2.23, p<×10^−4^, Fig. 6). There was significant heterogeneity (Q=37.28, p=0.0048) but no evidence of publication bias (p=0.45, Fig. 6). This effect remained significant when restricting studies to those using criteria-defined MS (OR 1.94, 95% CI 1.72-2.18, Fig. 6).

### EBV DNA detectable by PCR

31 full text papers were reviewed and 23 included in the analysis (supplementary references). 8 papers studied EBV DNA in CSF, 3 in whole blood, 7 in peripheral blood mononuclear cells, 4 in plasma/serum and 1 in saliva. The EBNA gene was the most commonly used for EBV detection (9 studies), with BAM used in 4 studies, VCA in 3 studies, and LMP in 2 studies.

EBV DNA was detectable in whole blood/PBMC more often in people with MS versus controls (n = 1853, 9 studies, OR 3.48, 95% CI 1.7360-6.9659, p<5×10^−4^). There was evidence of significant heterogeneity (Q=48.94, p<1×10^−4^) but no evidence of publication bias (p=0.78). Detection of EBV DNA did not differ between MS and control serum/plasma samples (n = 607, OR 1.81, 95% CI 0.77-4.26; p=0.18) or CSF (n = 802, OR 1.74, 95% CI 0.97-3.12, p = 0.062).

## Discussion and conclusions

There is a considerable body of epidemiological evidence implicating EBV in the pathogenesis of MS. EBV infection appears to be a necessary but not sufficient requirement for developing MS, EBV seroprevalence is higher among people with MS, symptomatic EBV infection (IM) is more prevalent among people with MS, and the effect of anti-EBV antibody titre on MS risk appears contingent on the strongest genetic risk factor for MS, the HLA-DRB1*1501 genotype.

In our meta-analysis of interaction between EBV and other risk factors, we demonstrate evidence for supra-additive interaction between EBNA titre and HLA status in determining risk. The absence of strong evidence for interaction between EBV and other risk factors in our analysis demonstrates the importance of using multiple measures of interaction (AP, RERI, Synergy Index, and multiplicative interaction) to avoid the risk of type 1 error. Relying on the Attributable Proportion alone to make inferences about interaction may be misleading. However, the small number of studies suitable for our analysis of interaction limits the power of this meta-analysis, and therefore conclusions about interaction should be drawn cautiously from these results.

The mechanism via which EBV exerts this increased risk remains unknown, and our systematic review of the literature highlights a multitude of potential biological mechanisms that have been both demonstrated, replicated, and importantly not replicated. It seems likely that the route via which EBV exerts its effect lies in complex interactions between EBV and the host genome, the precise mechanisms of which remain to be elucidated. Despite the relatively small retrospective studies that have been performed, it also appears clear that IgG titres against EBV antigens are not able to act as a clinically useful biomarker in either treated or untreated cohorts.

Our results for the seroprevalence of EBV among people with MS are consistent with the previously published meta-analysis, which reported ORs of 4.47 (95%CI 3.26-6.11) and 4.51 (95%CI 2.84-7.16) for EBNA and VCA respectively. Our estimates of 3.39 (95%CI 2.53-4.56) and 3.18 (95%CI 1.88-5.38) are more conservative, likely reflecting new, larger studies with smaller effect sizes and our different inclusion criteria^7^. Similarly, our estimates of measures of interaction between EBNA titre, HLA status, and MS risk are similar, though not identical, to the published meta-analysis estimates^35^. The reasons for this discrepancy are not clear.

An EBV vaccine has previously been developed, which prevents IM but not EBV seroconversion ^36^. Newer vaccines are entering phase I clinical studies. The data above indicates that IM carries an odds ratio of 5.11 in individuals with a high genetic risk - preventing IM in this population has the potential to significantly reduce MS risk. Approximately 50% of the general population are EBV seronegative at age 15 ^37^; hence vaccination in early adolescence to prevent IM seems an appropriate route both reduce IM-associated morbidity and subsequent EBV-related disease.

Despite the evidence above, not all epidemiological aspects of MS can be explained by EBV infection. The relatively short latency between putative infection and subsequent MS seen in the Faroe epidemics, and the decreasing risk in migrants moving from high-to low-risk areas cannot be explained purely by EBV infection - the fact remains that MS is overwhelmingly likely to be the result of multiple environmental risk modifiers. However, evidence for EBV infection as an obligate step in MS development is increasing, and with vaccination on the horizon as a potential preventive intervention, cannot be ignored.

## Data Availability

All data are publicly available

---

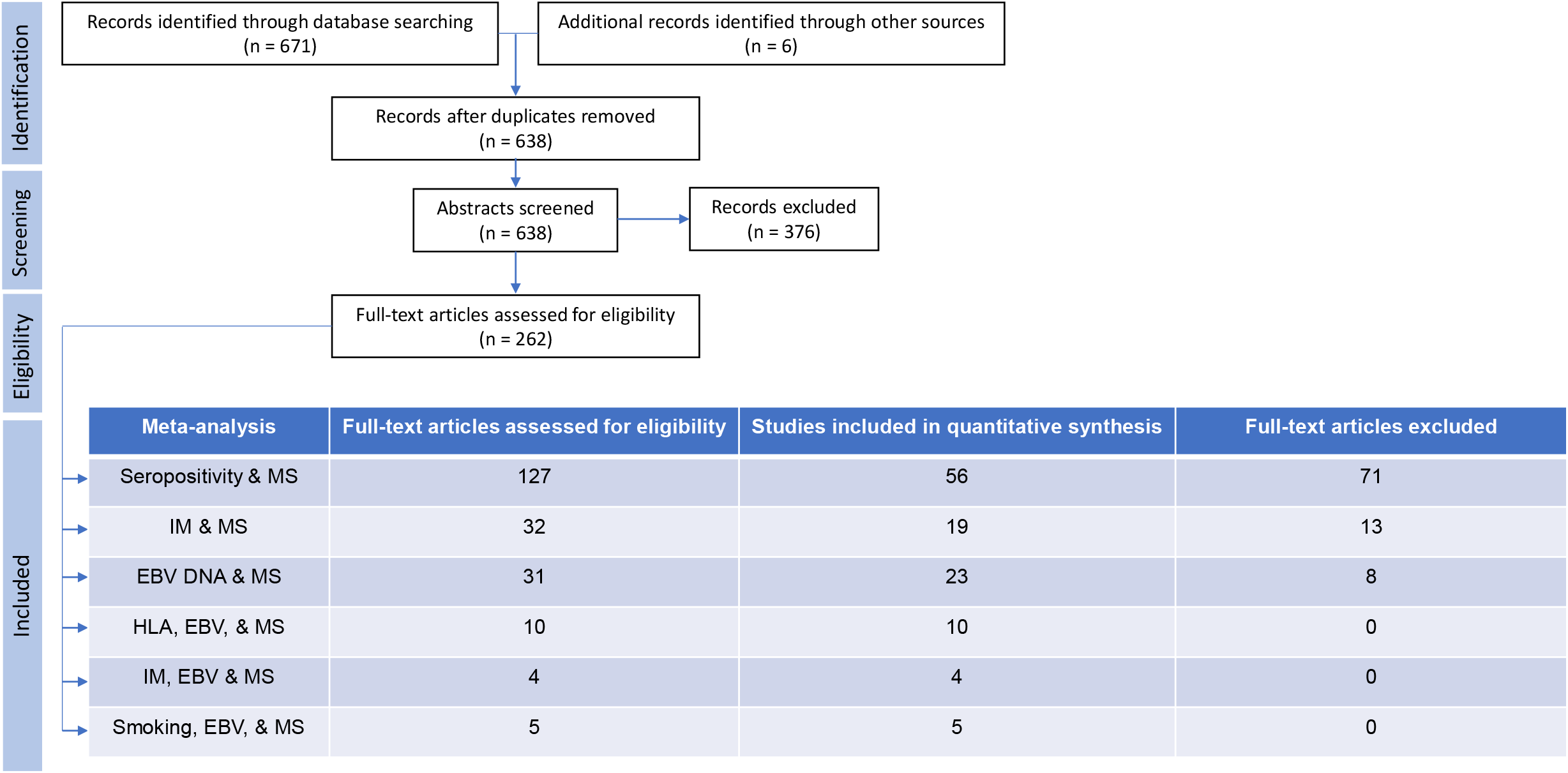

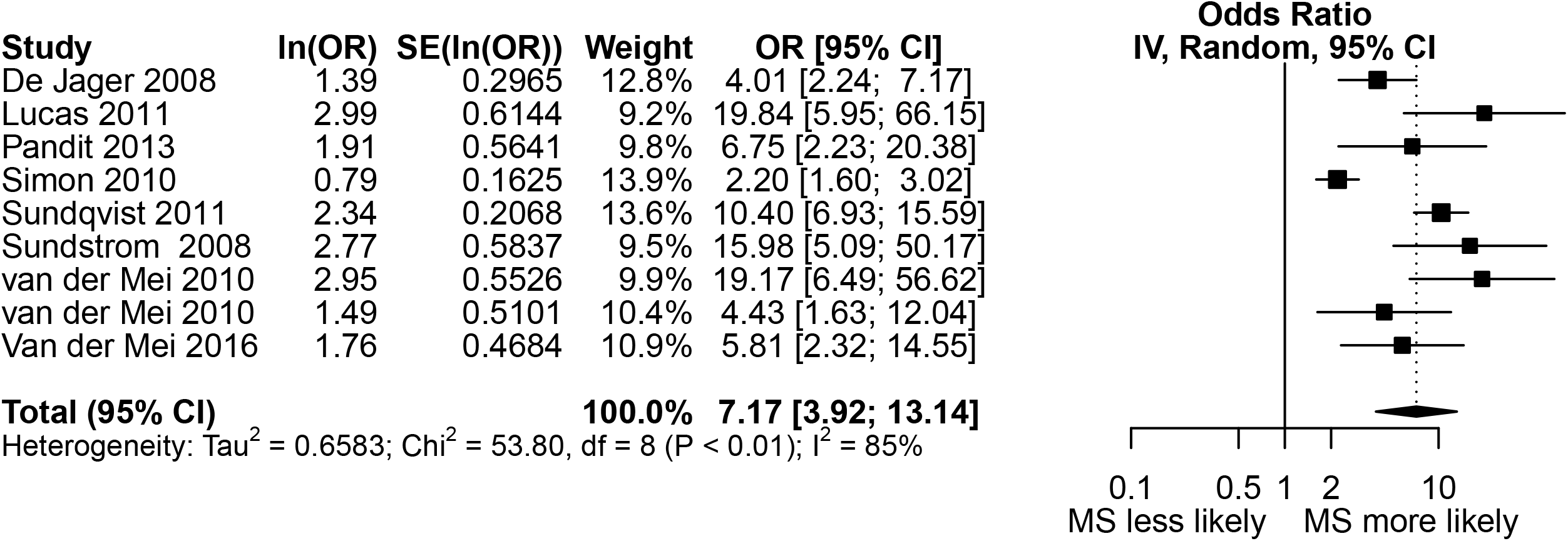

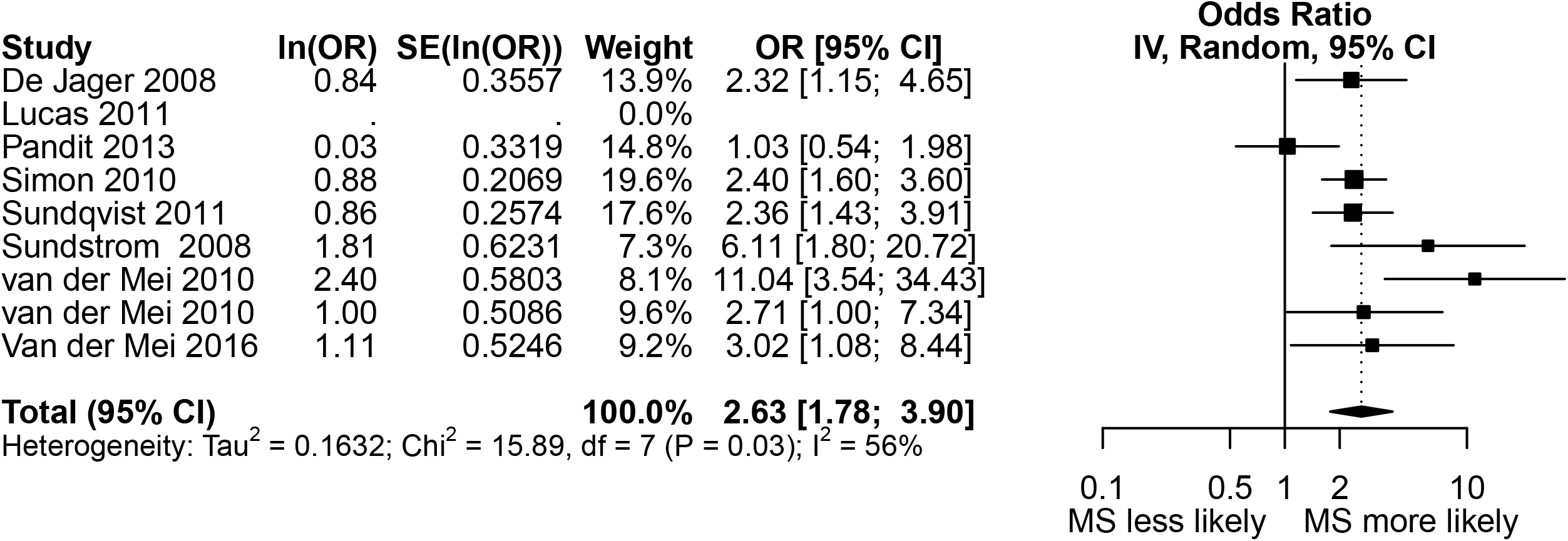

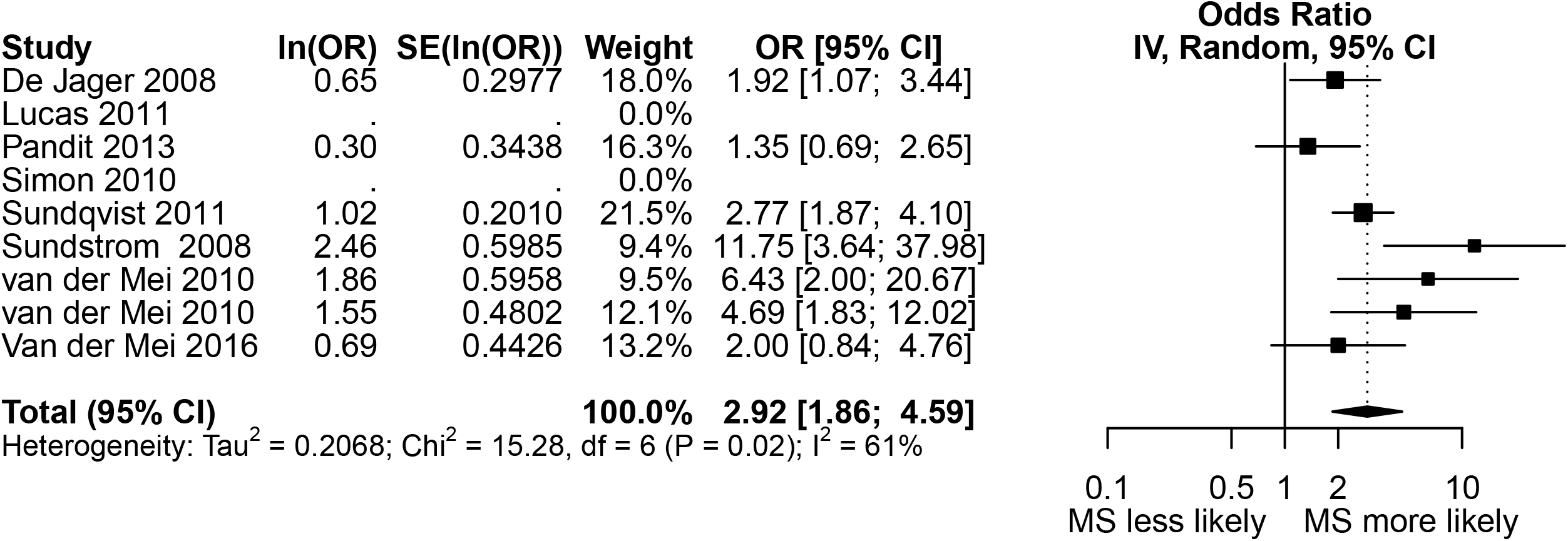

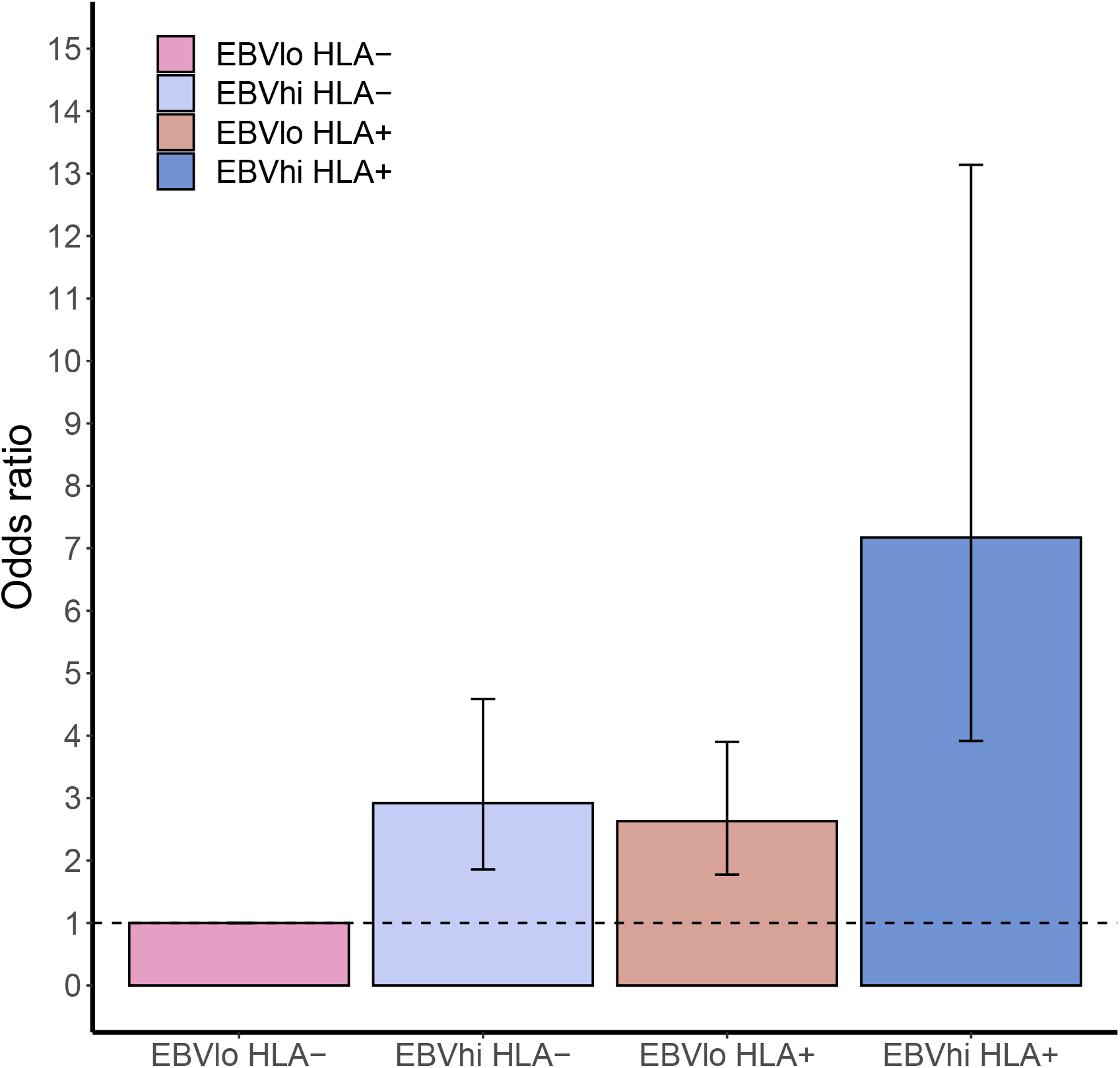

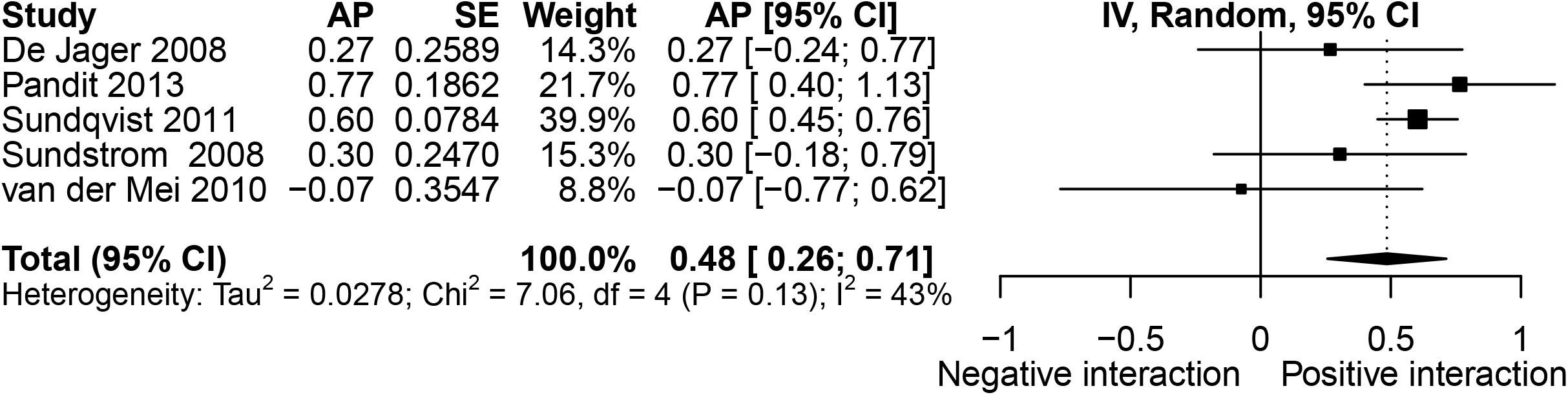

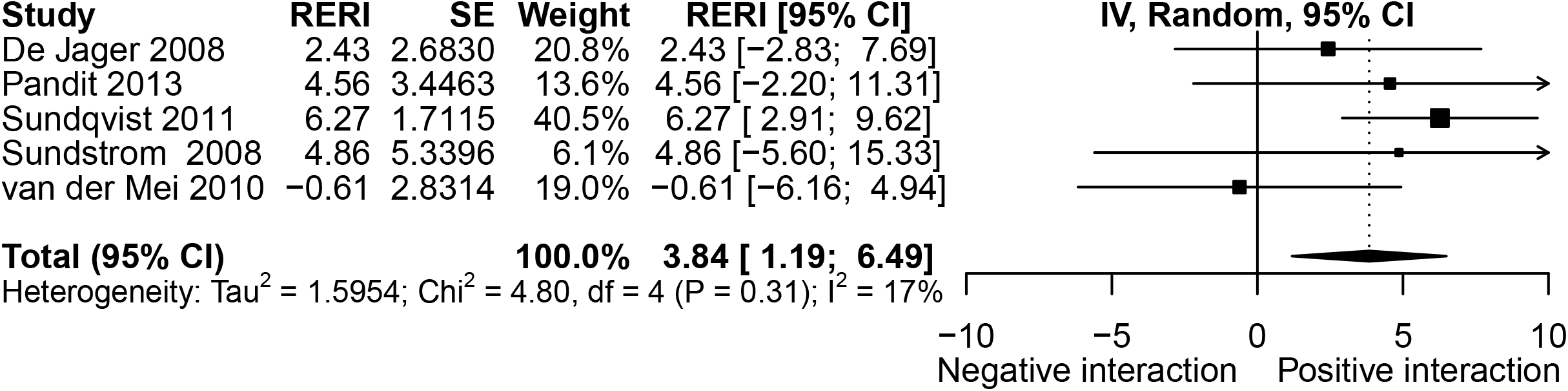

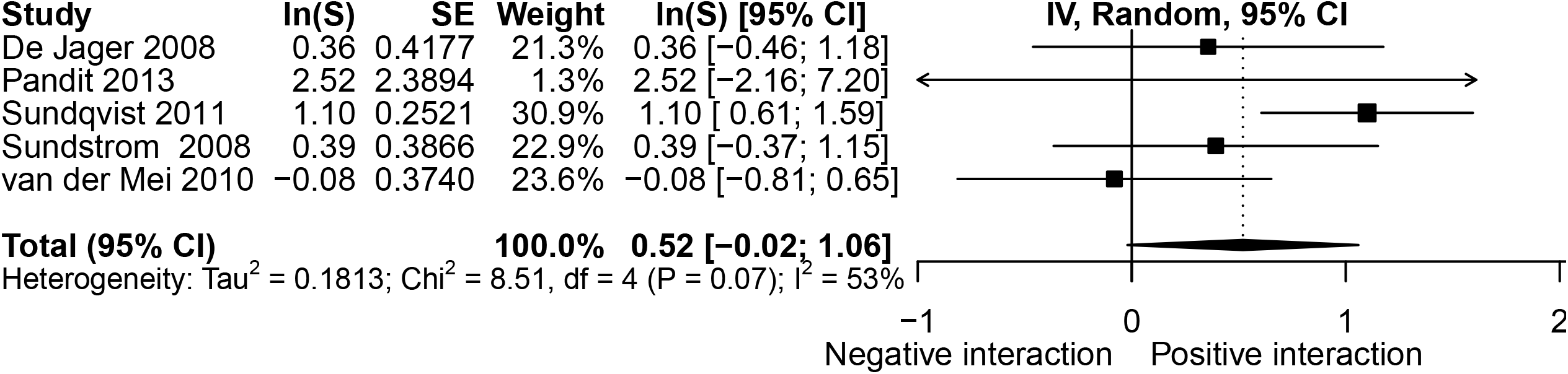

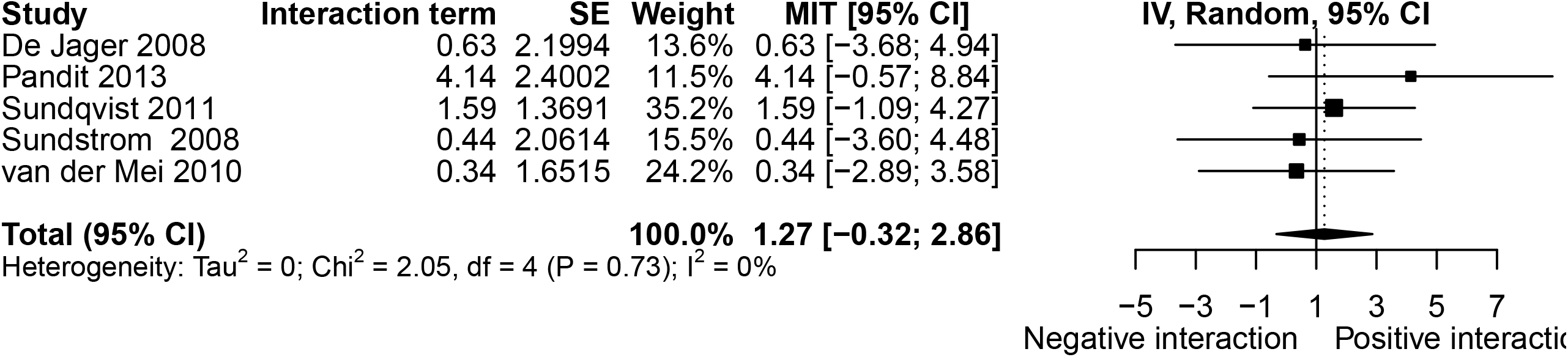

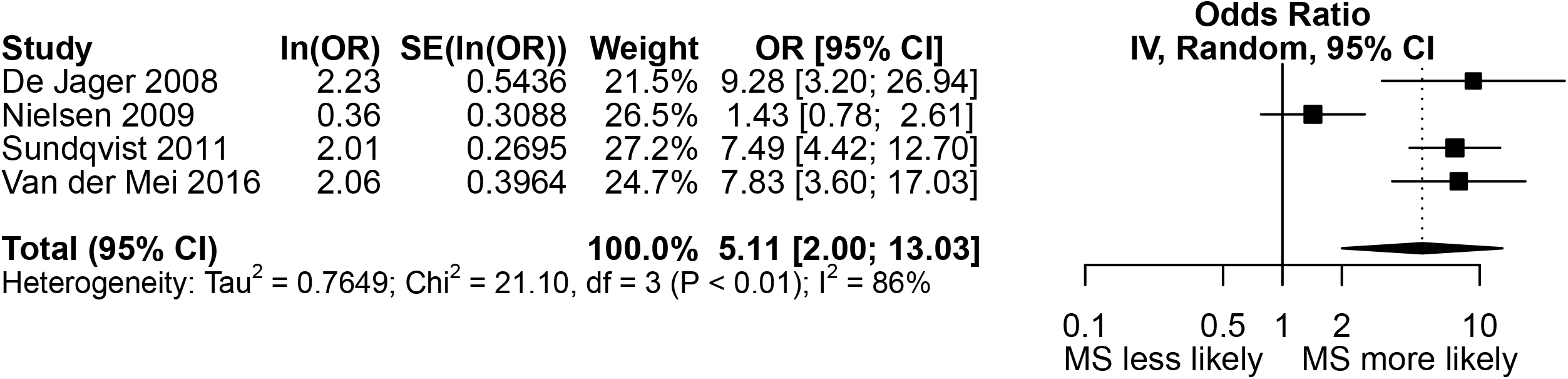

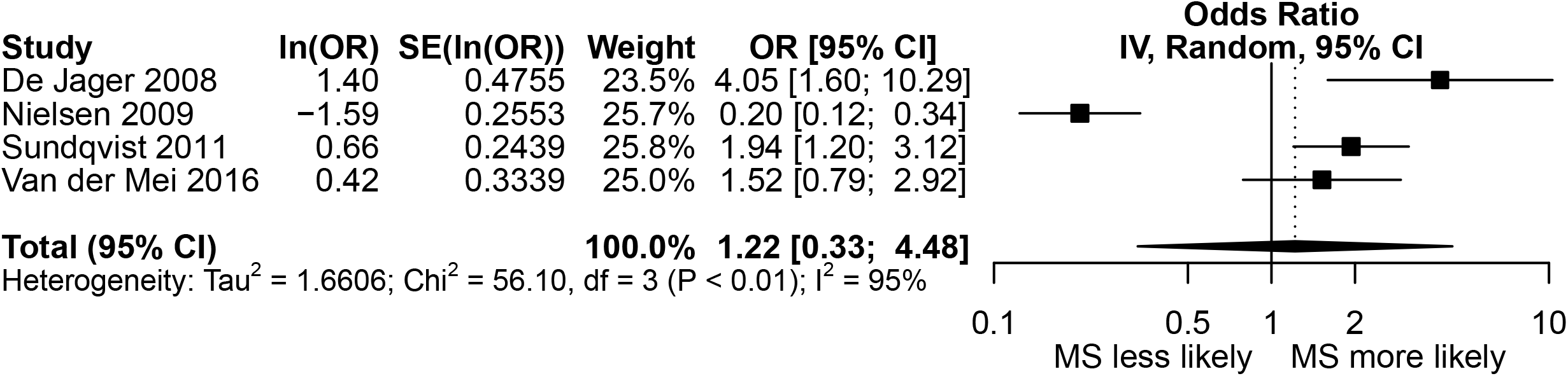

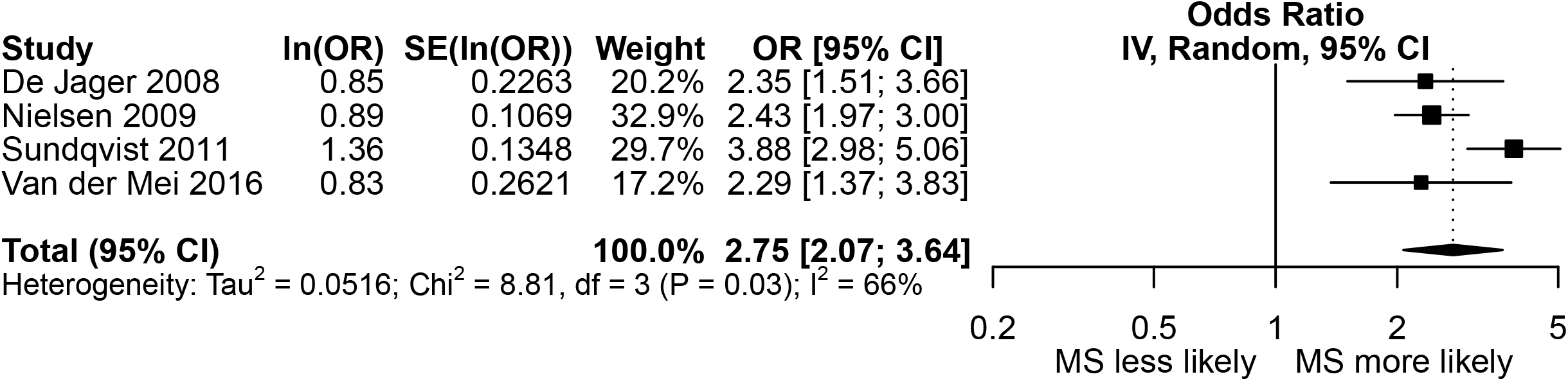

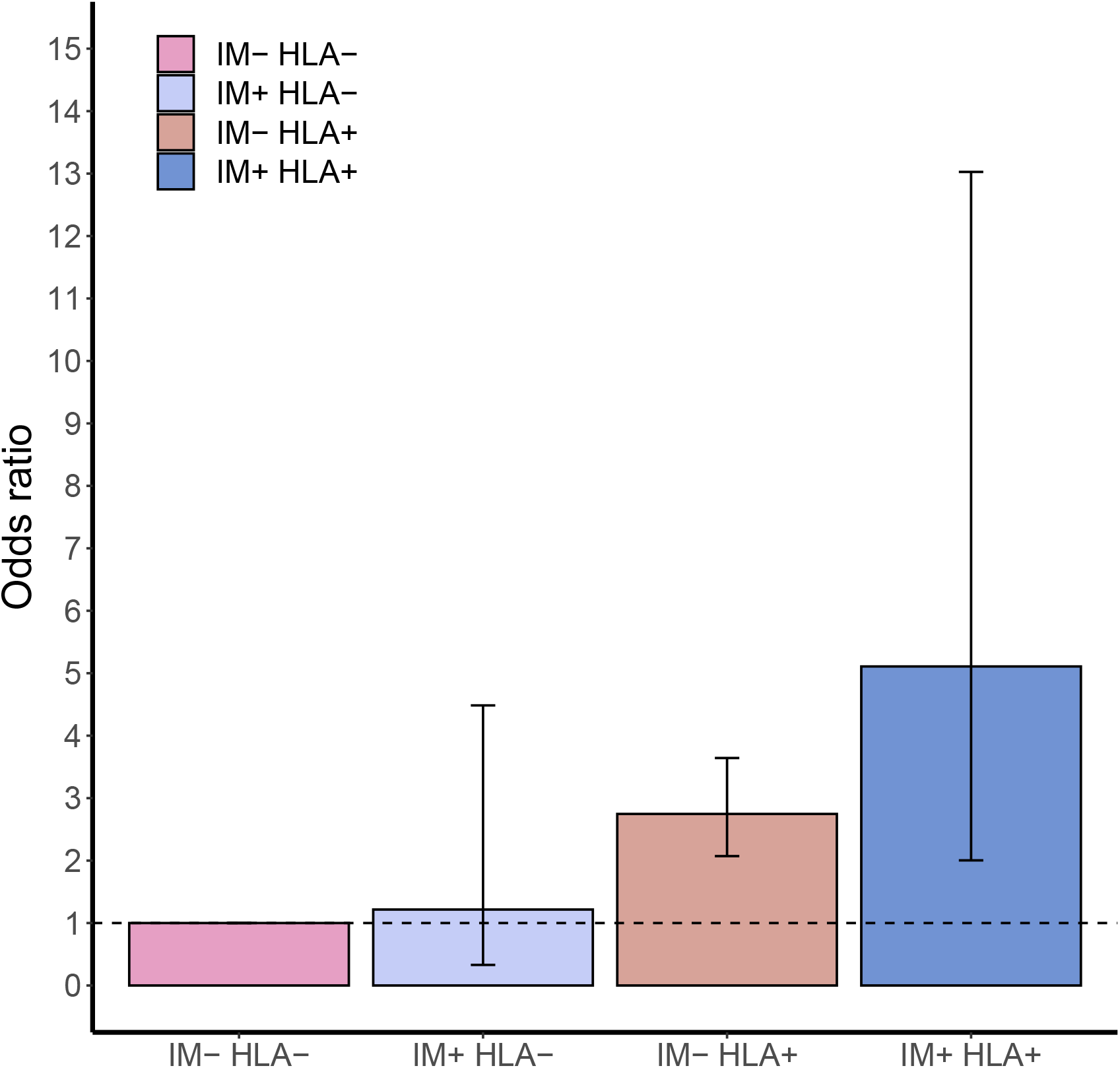

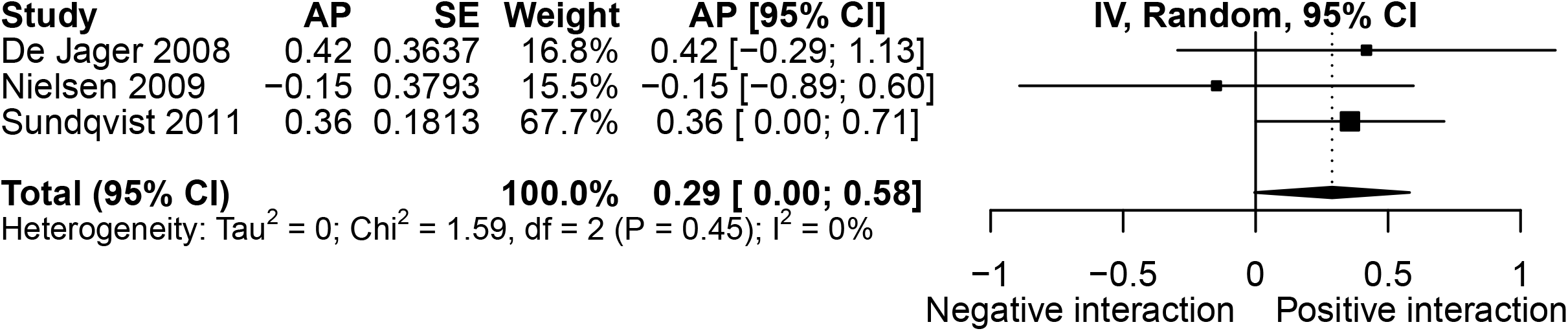

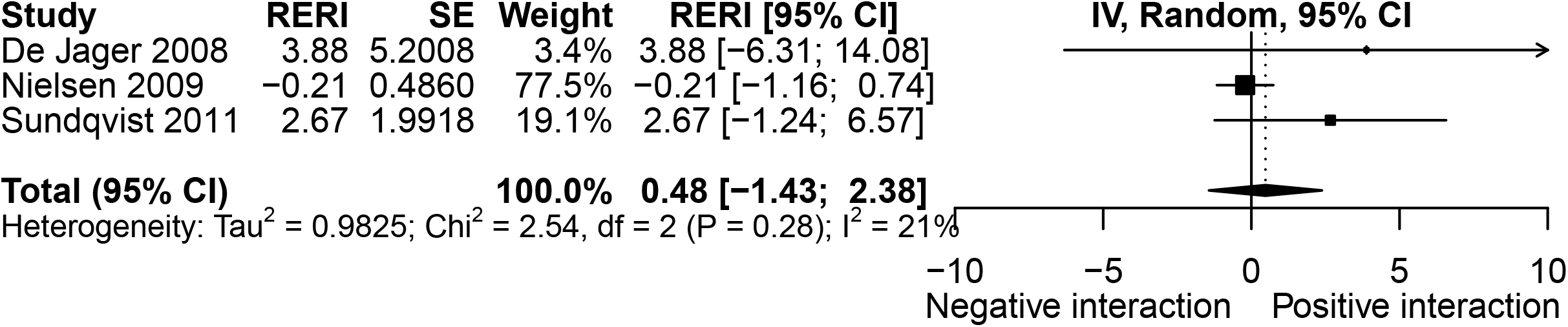

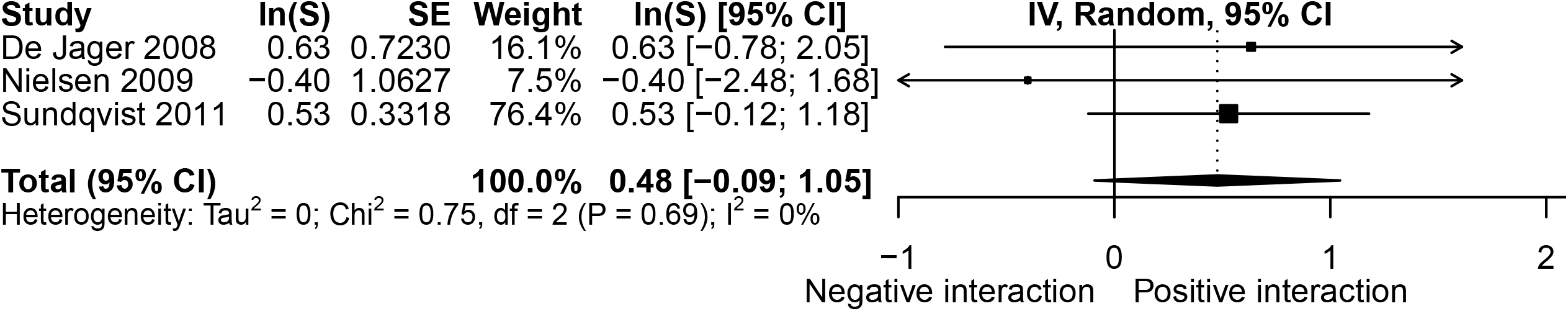

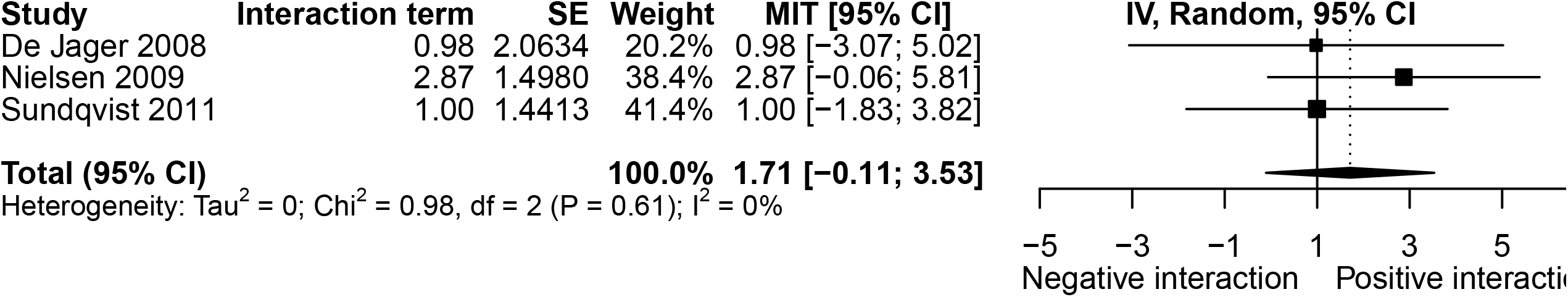

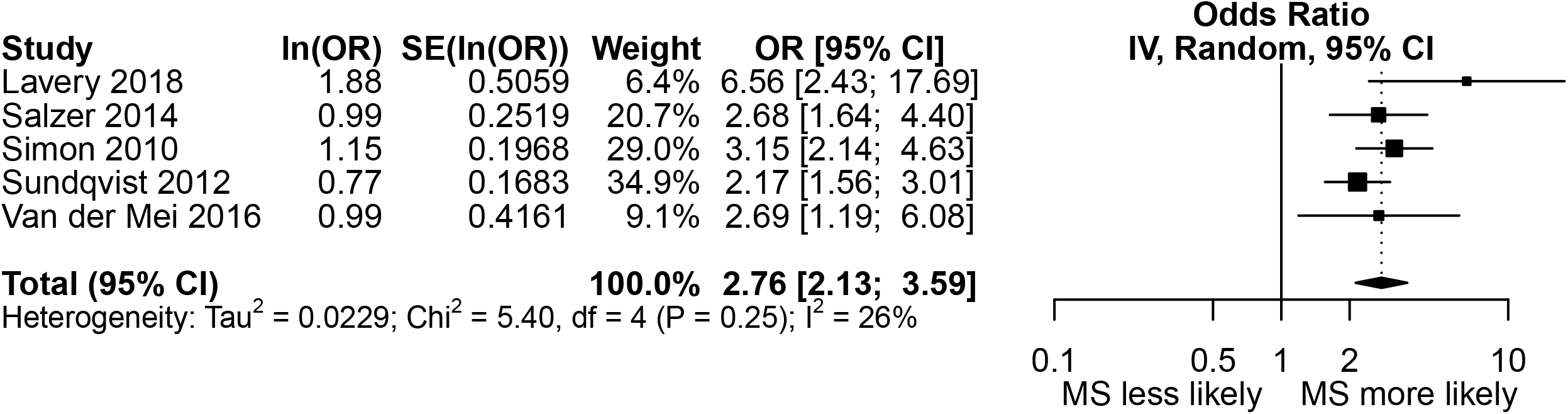

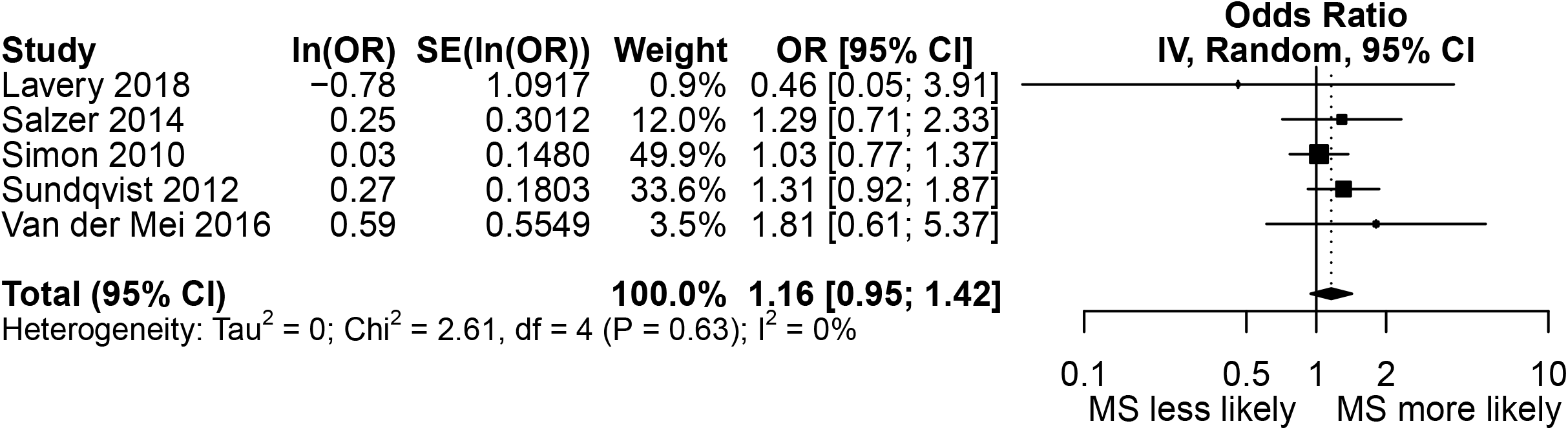

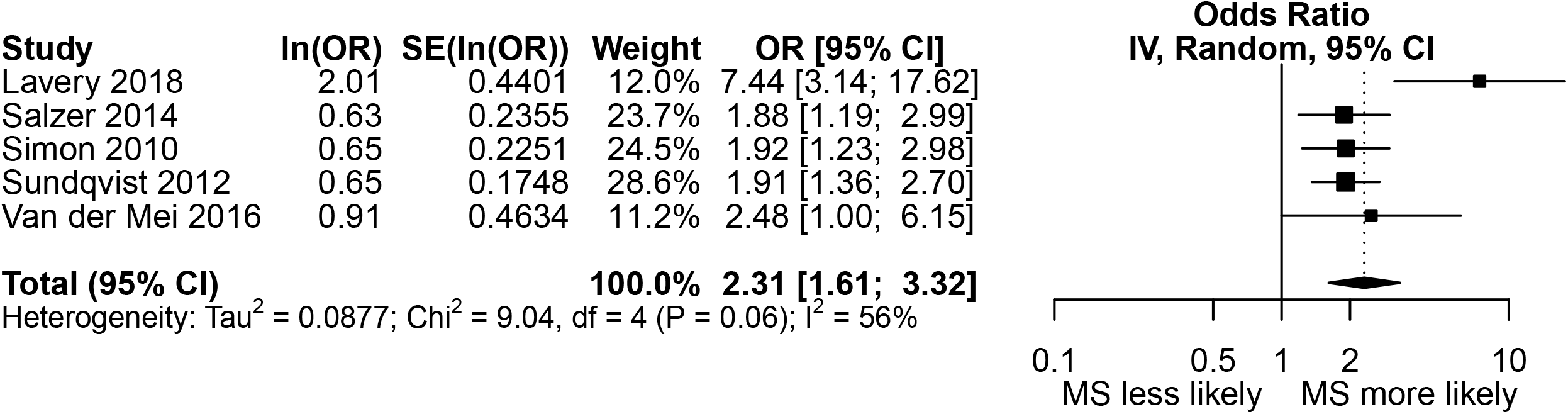

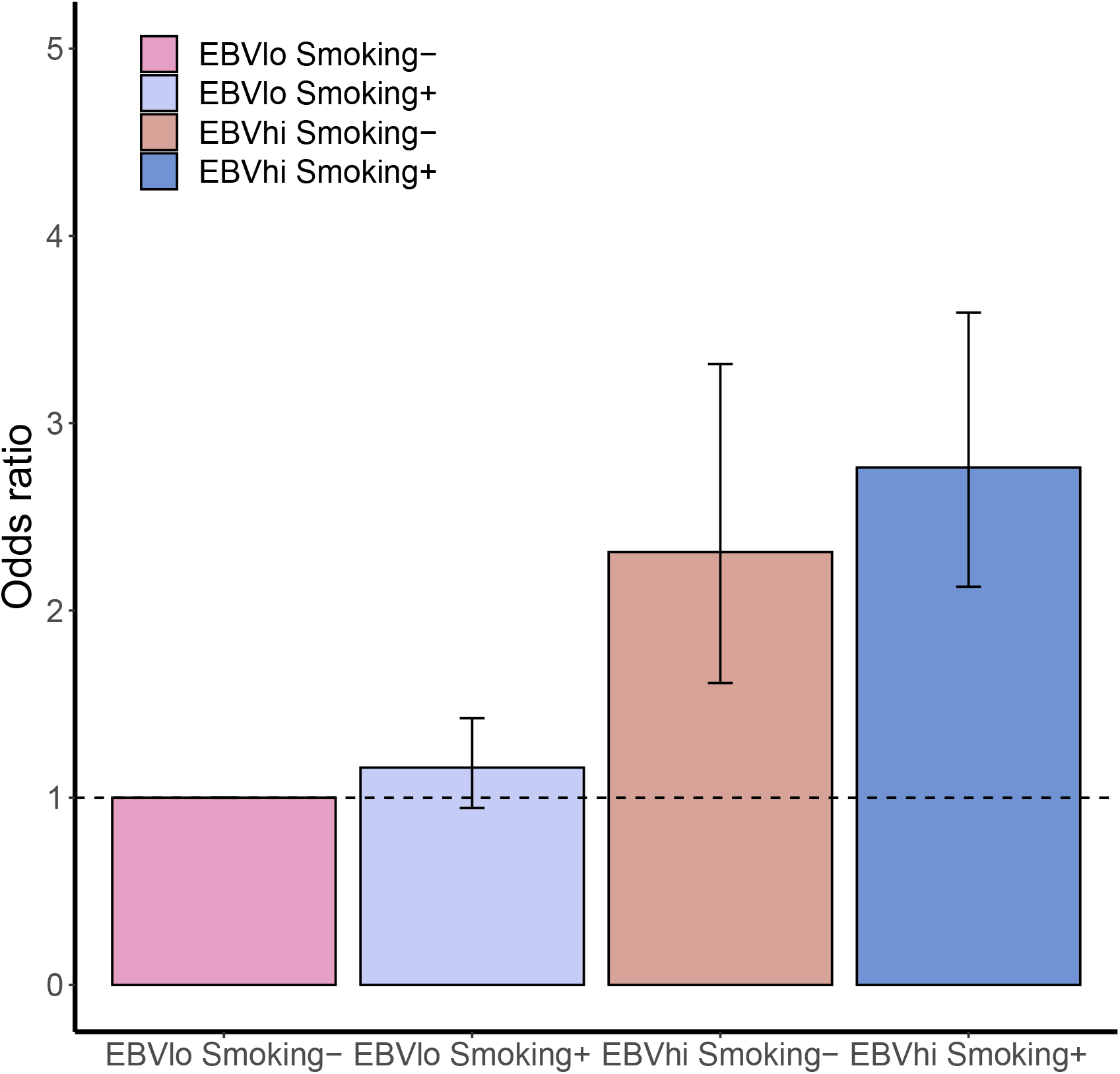

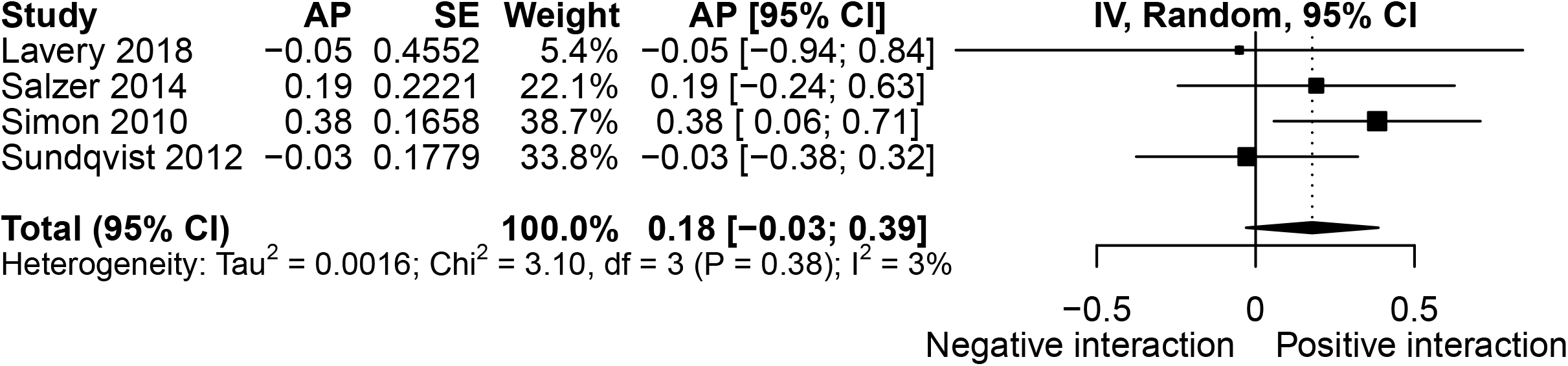

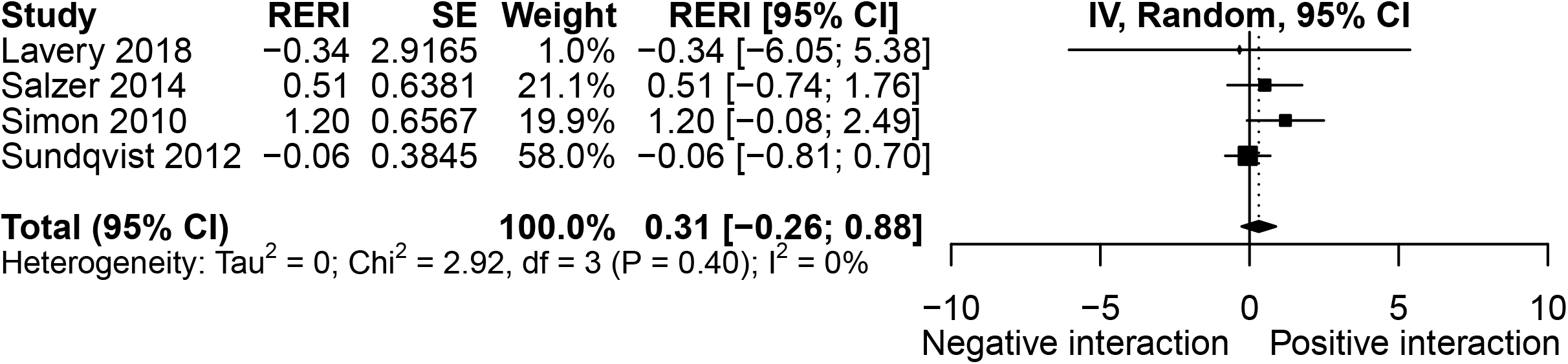

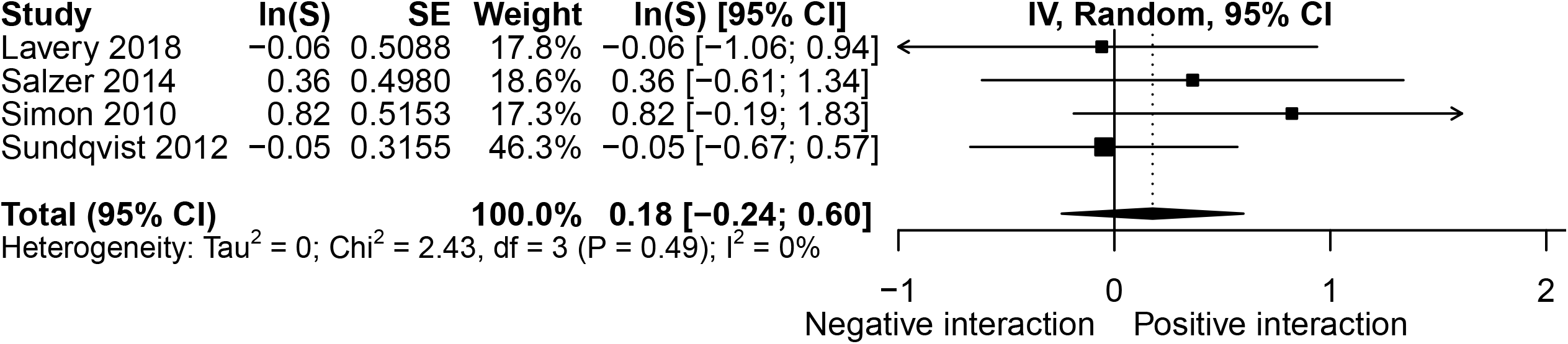

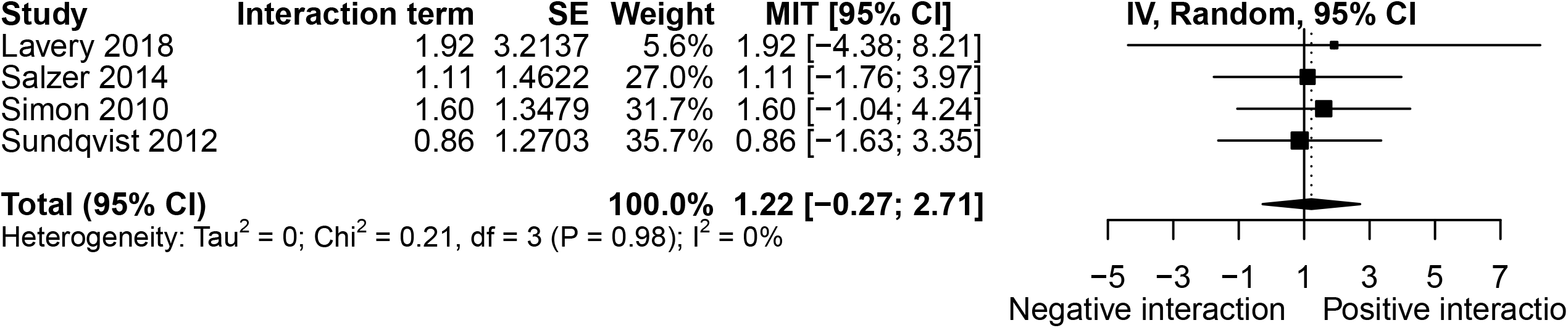

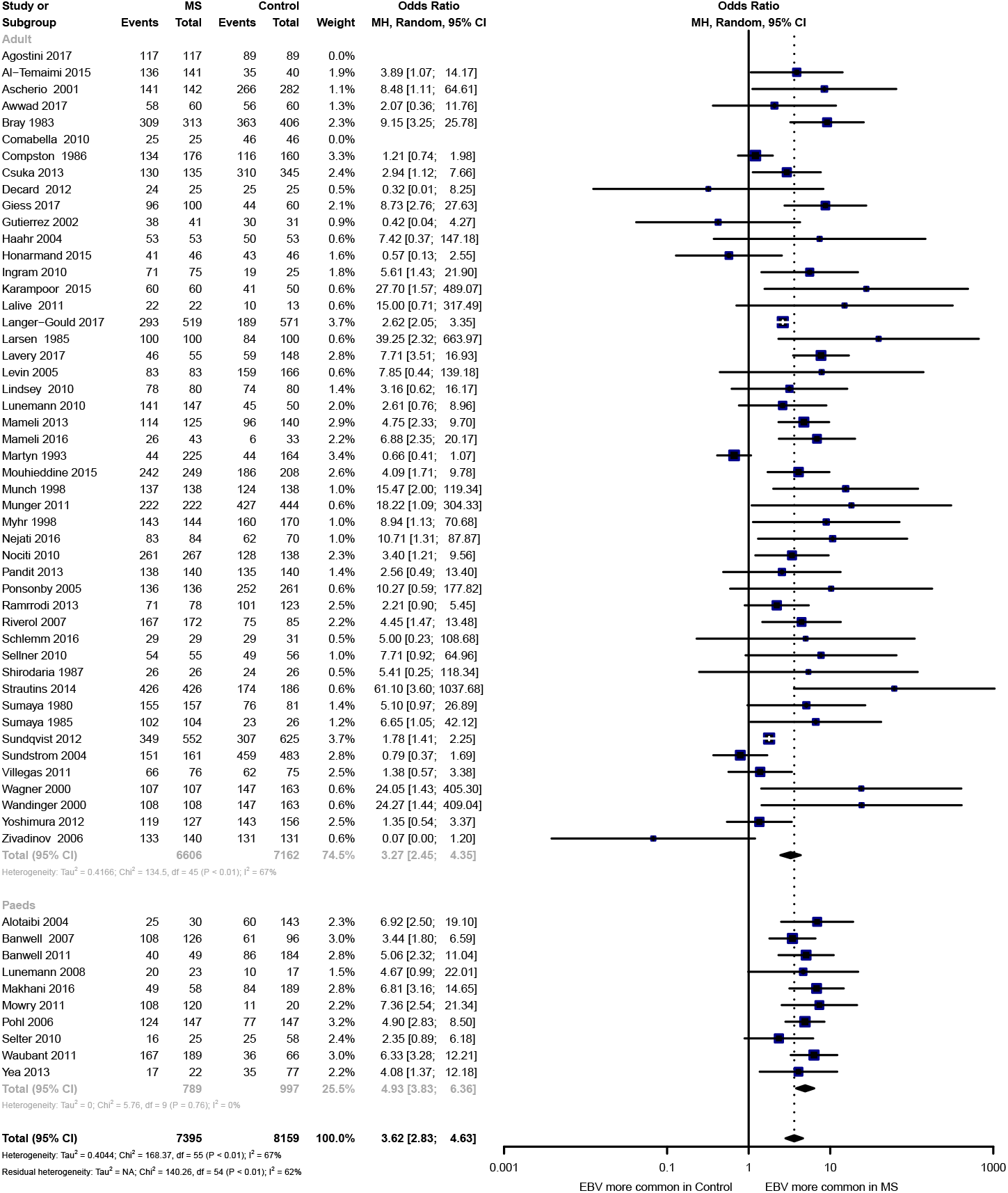

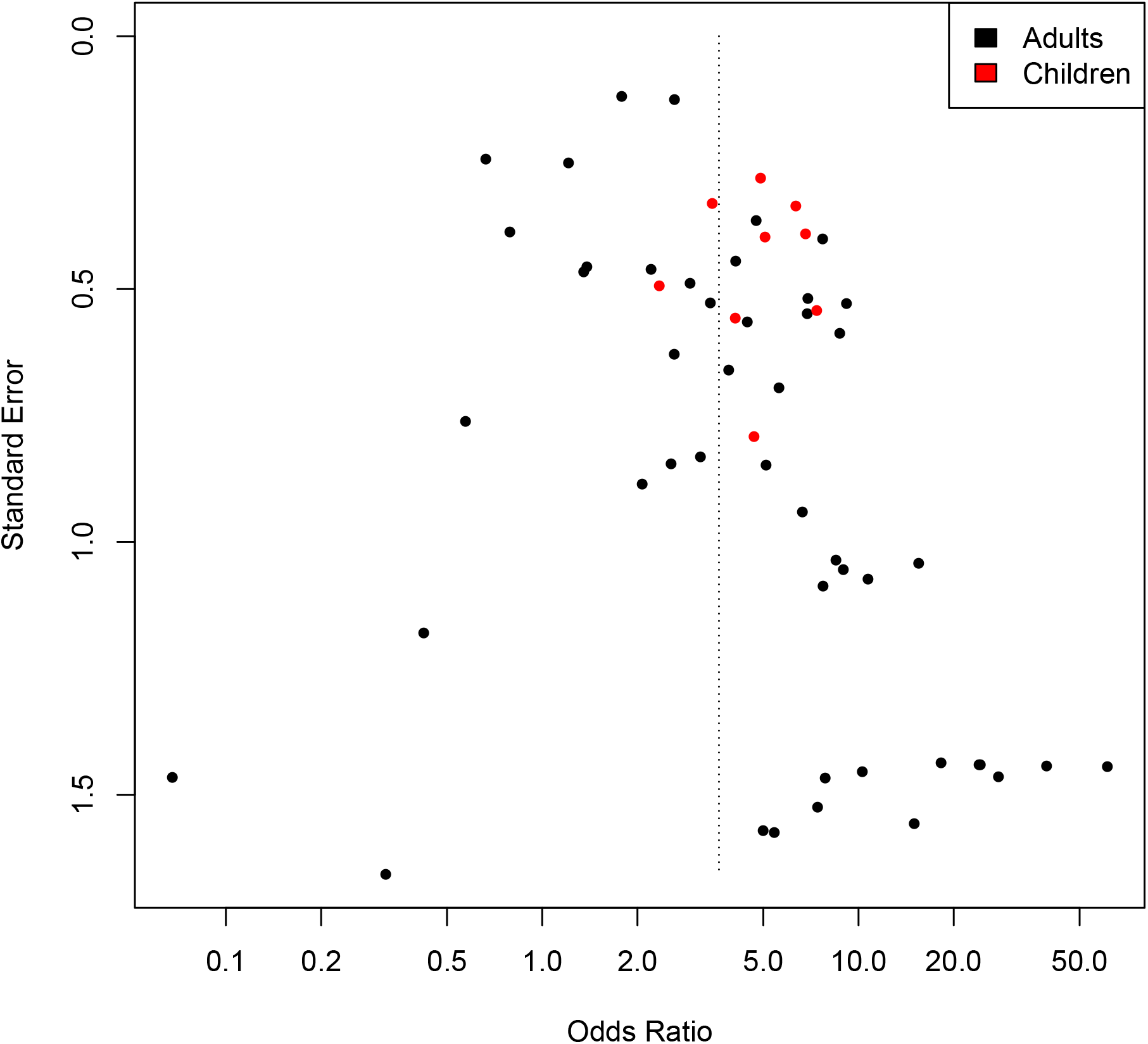

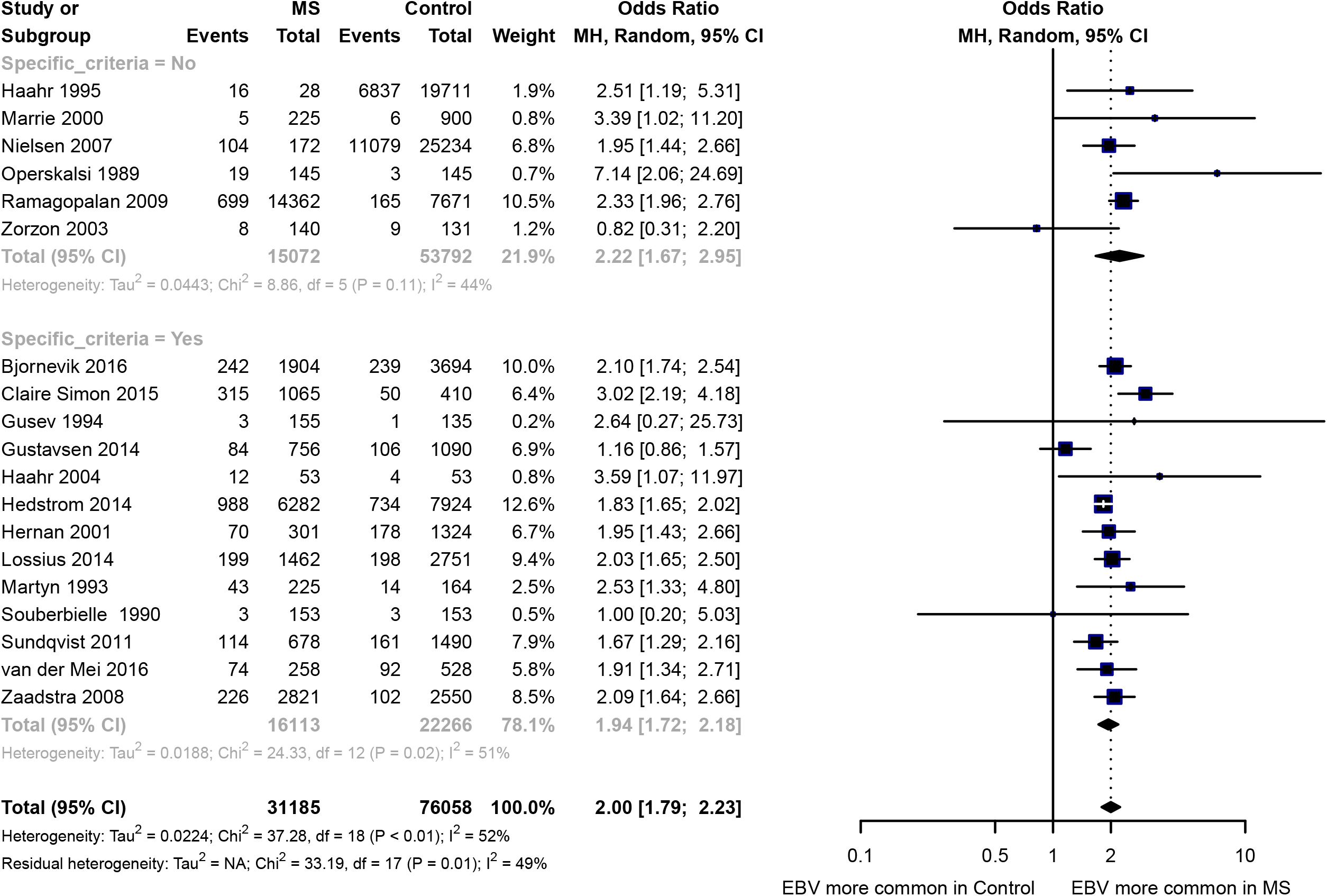

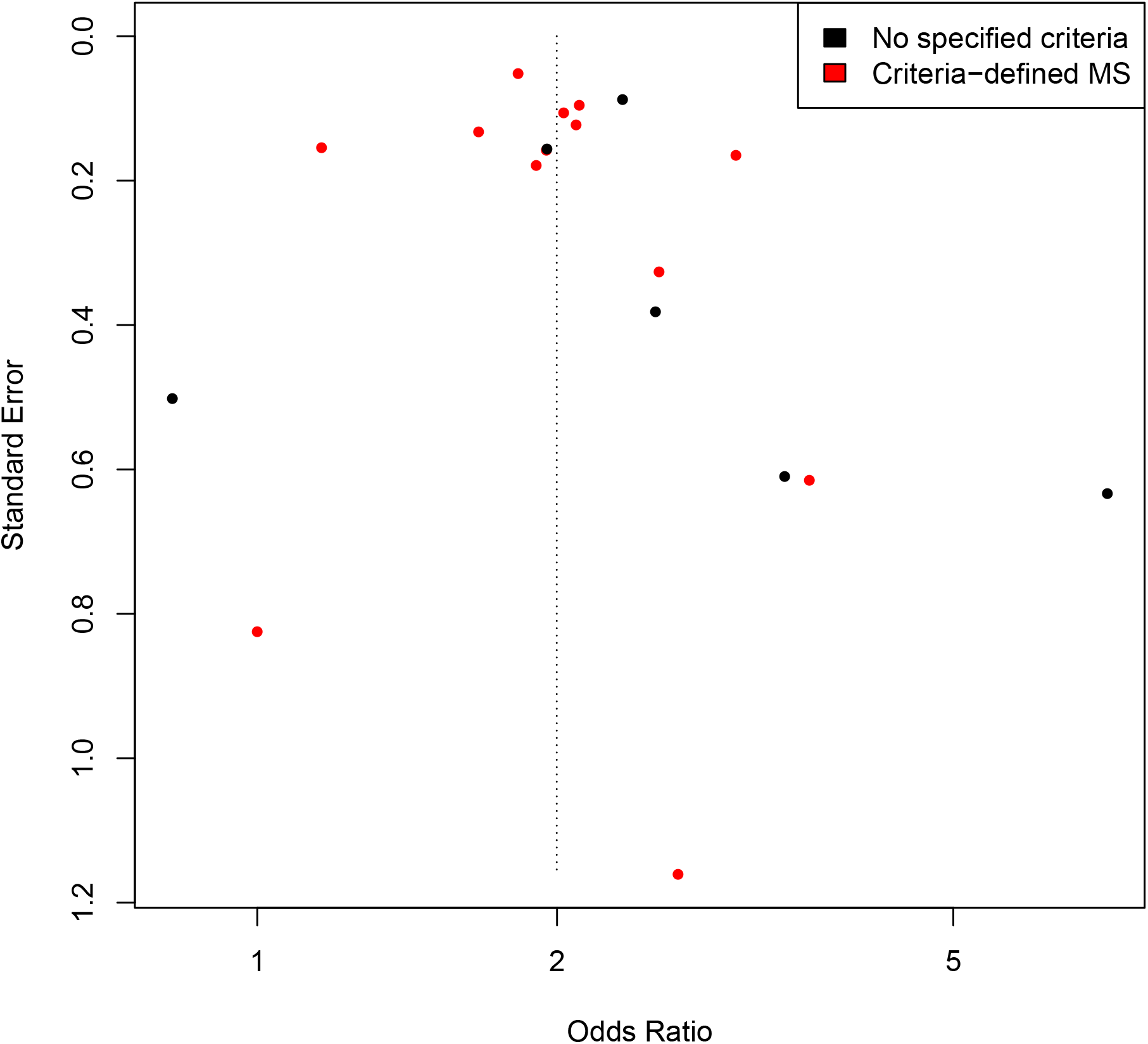

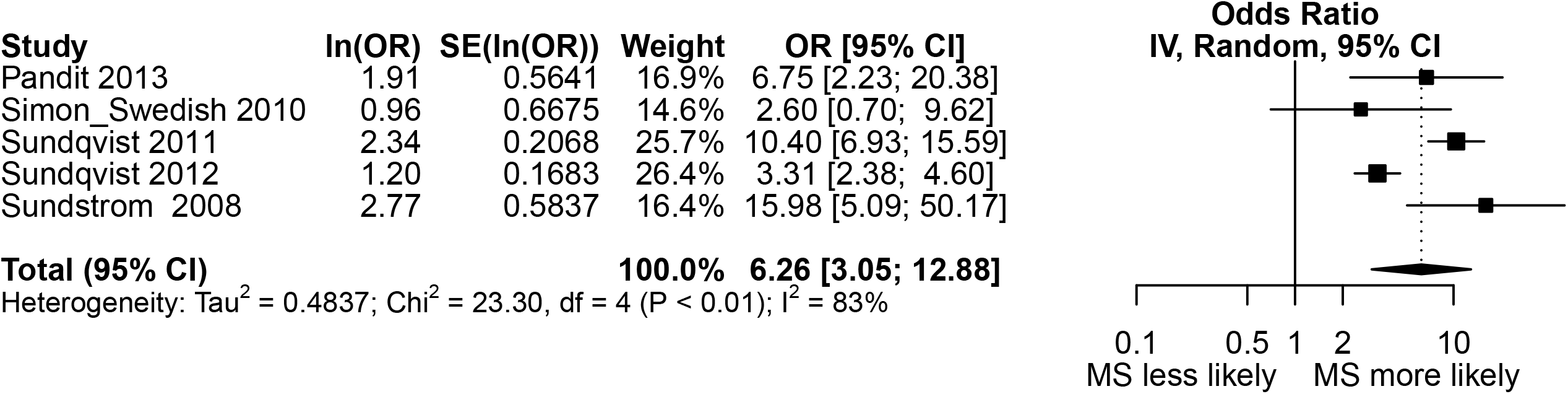

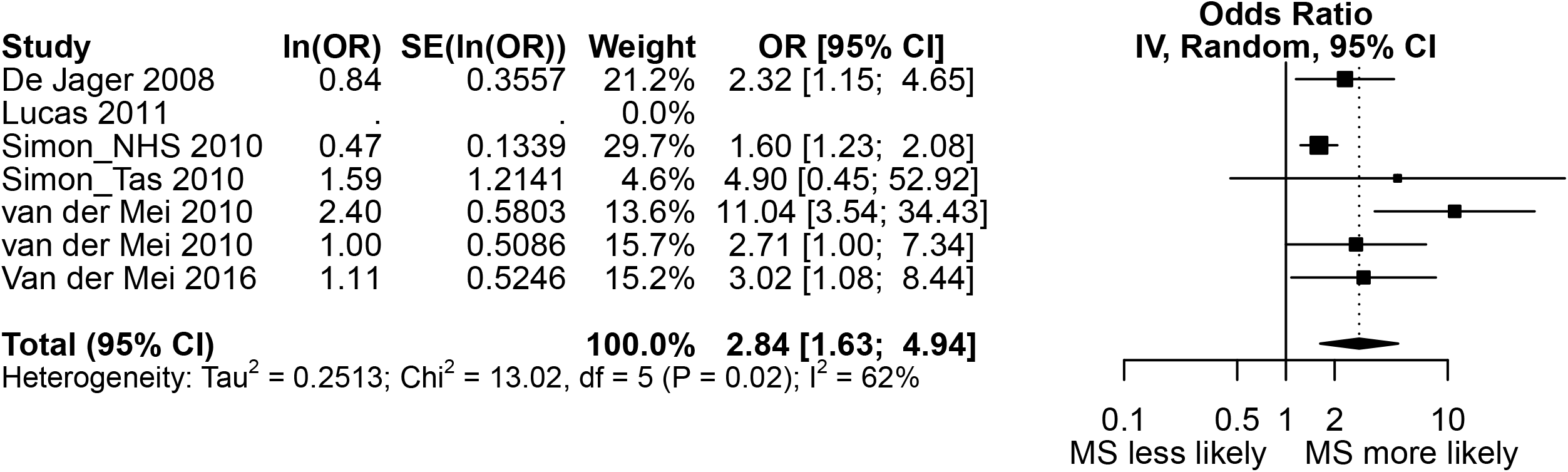

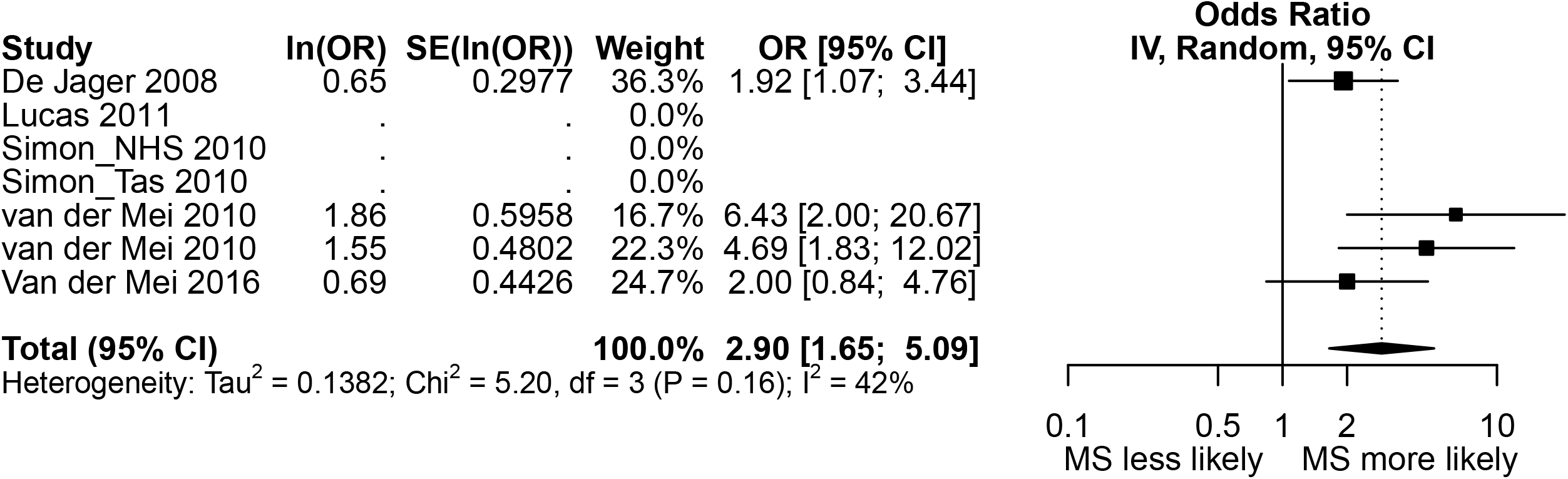

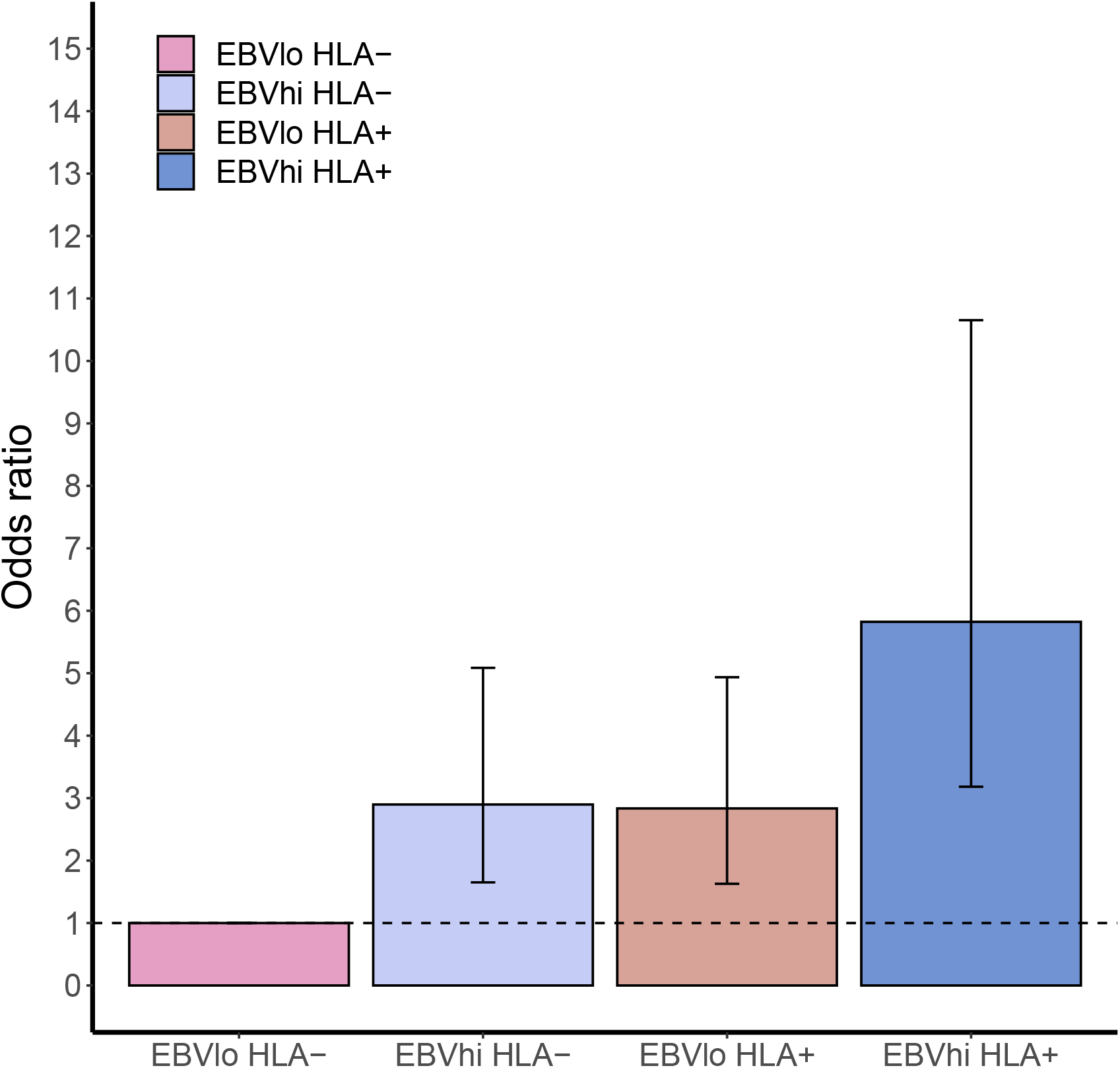

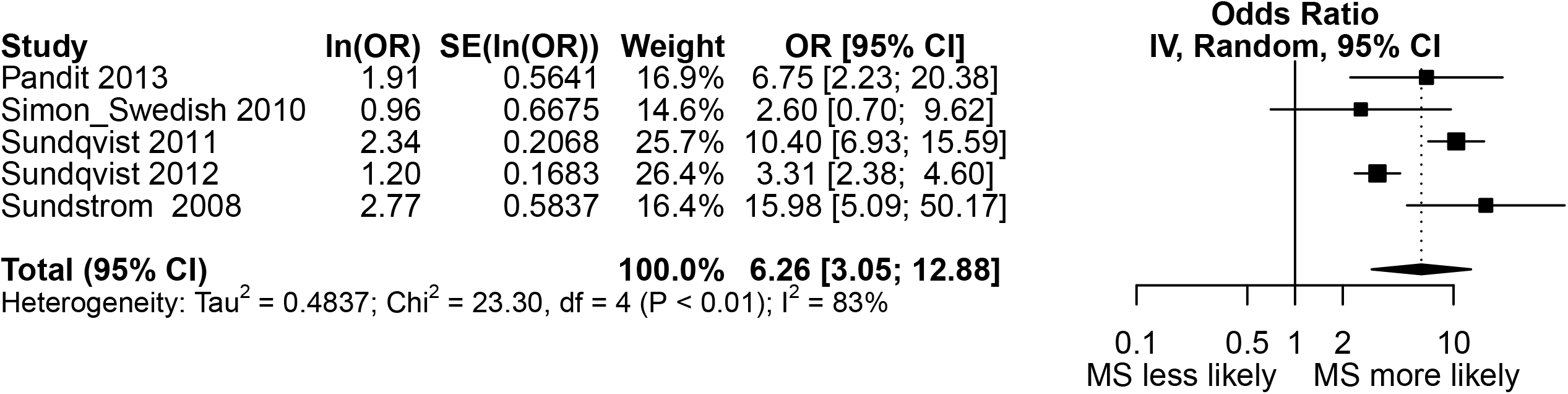

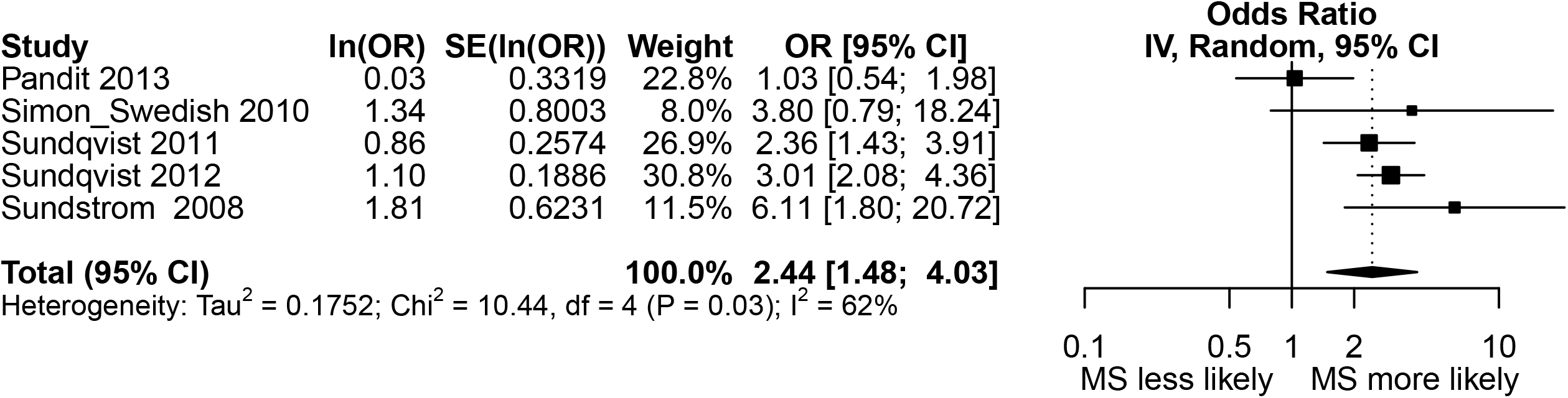

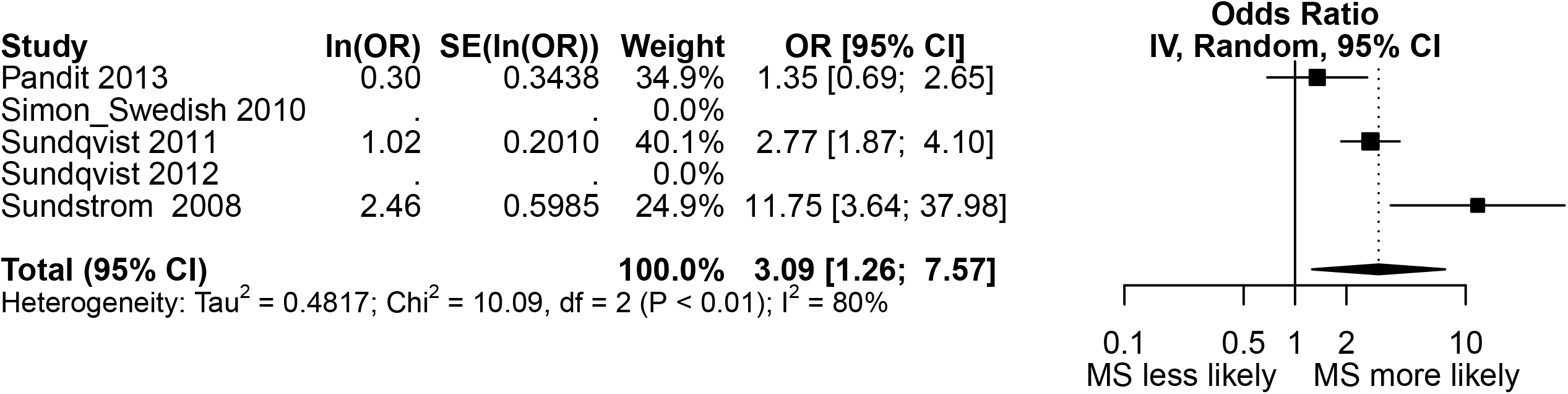

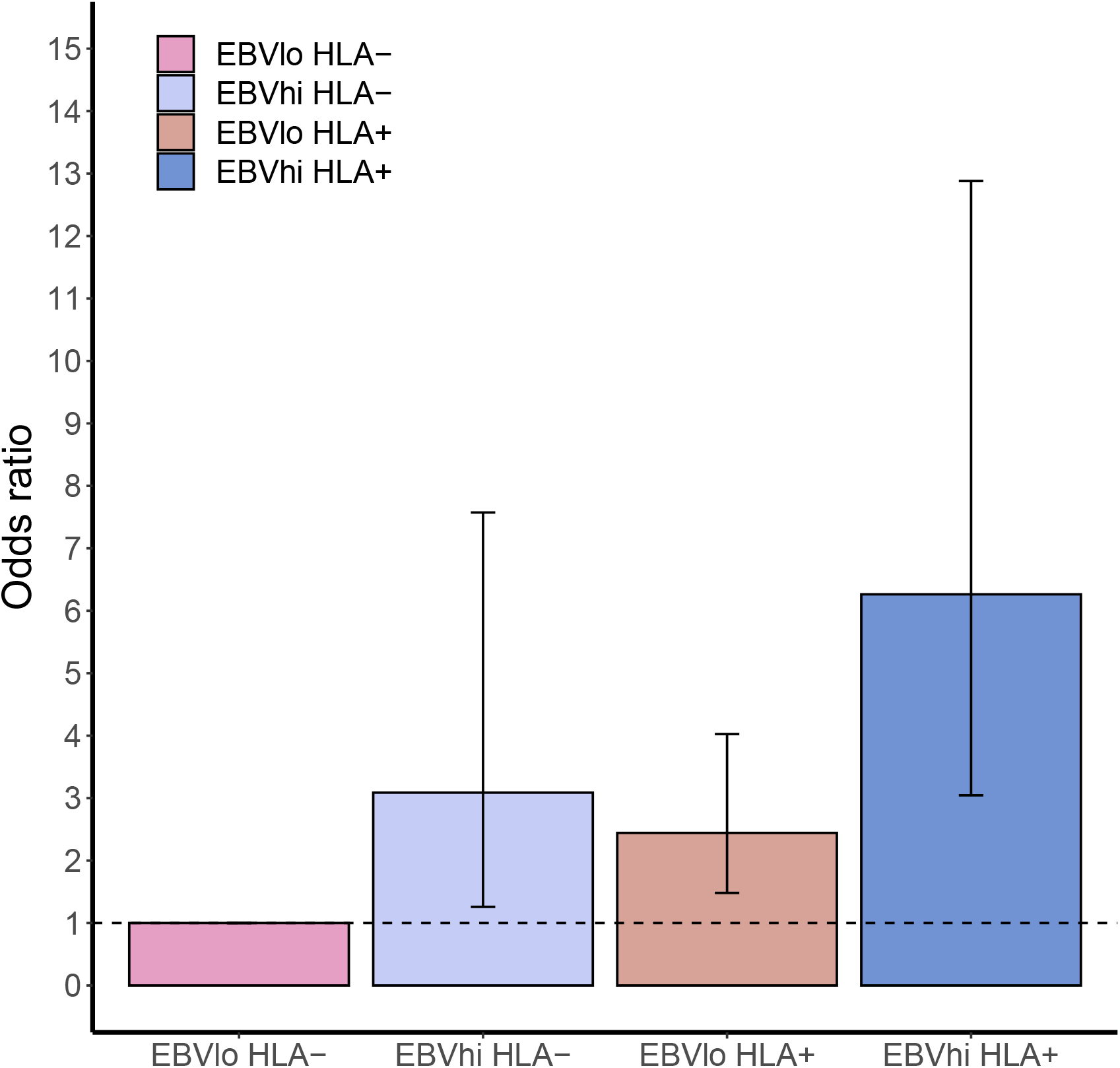

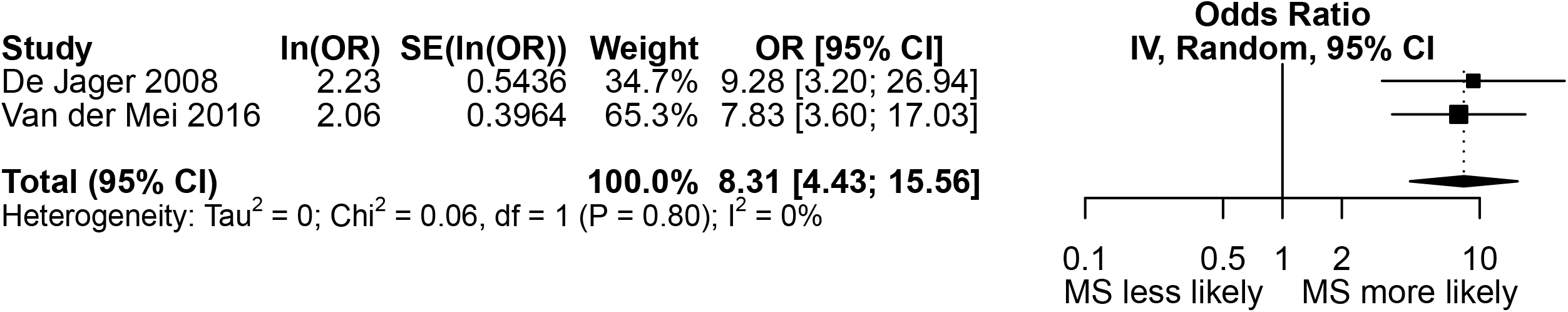

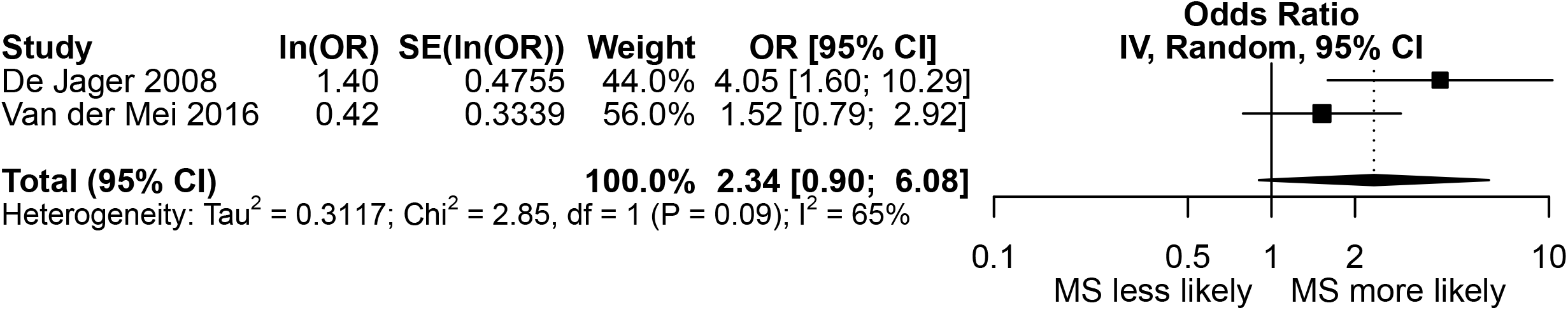

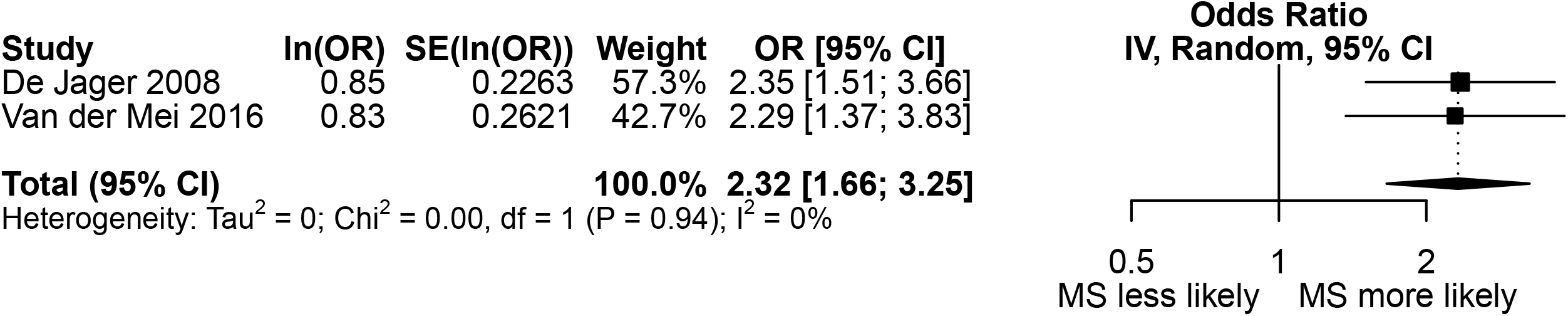

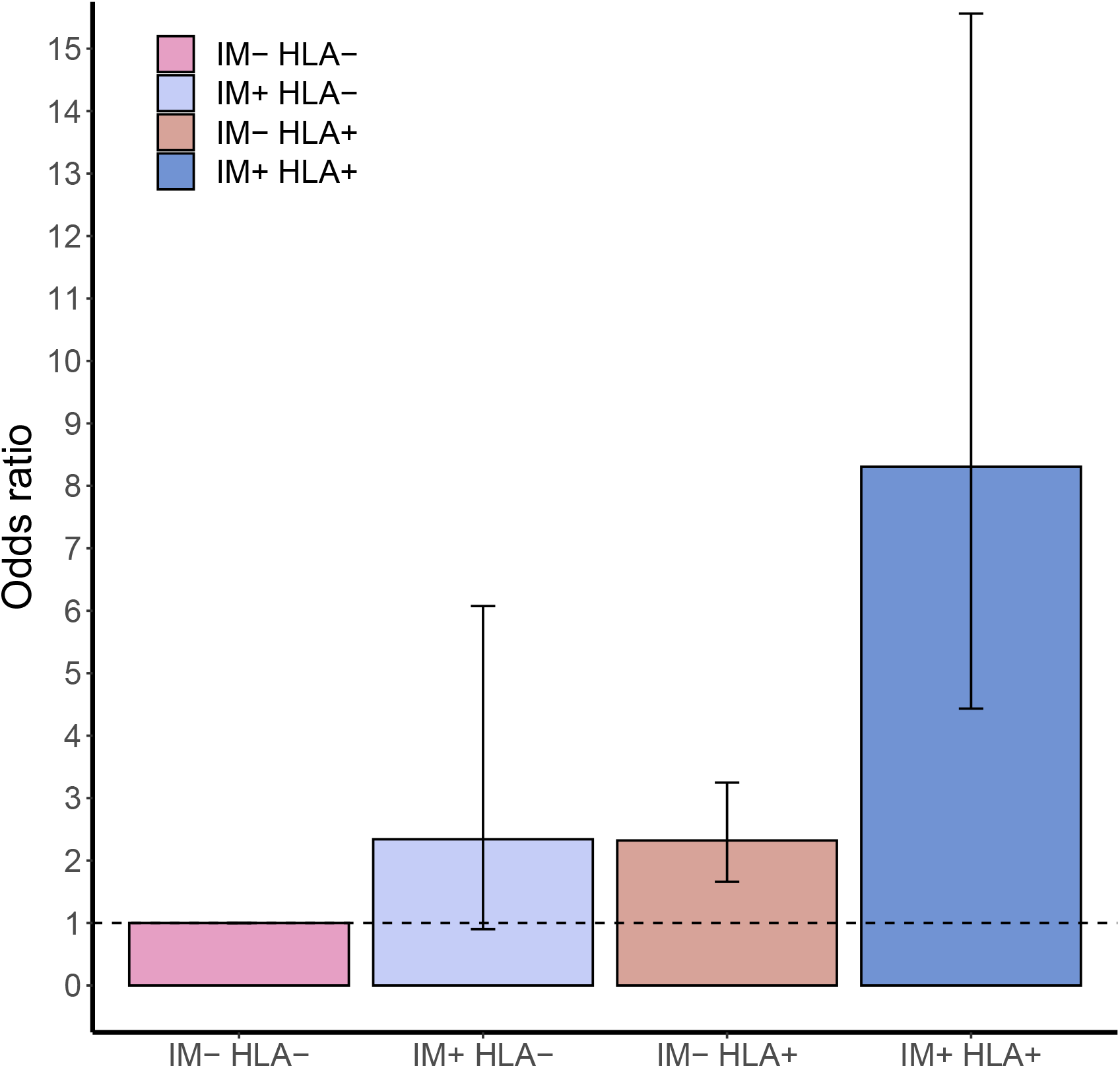

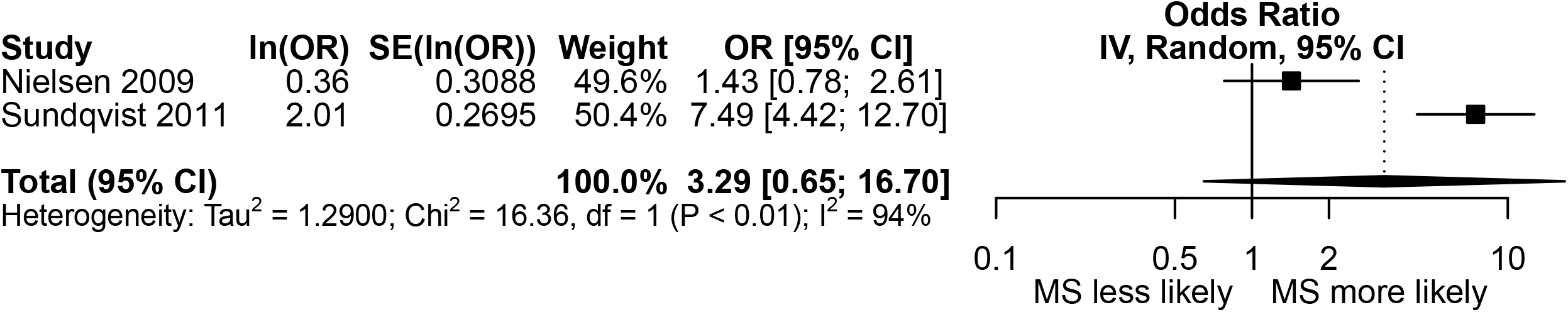

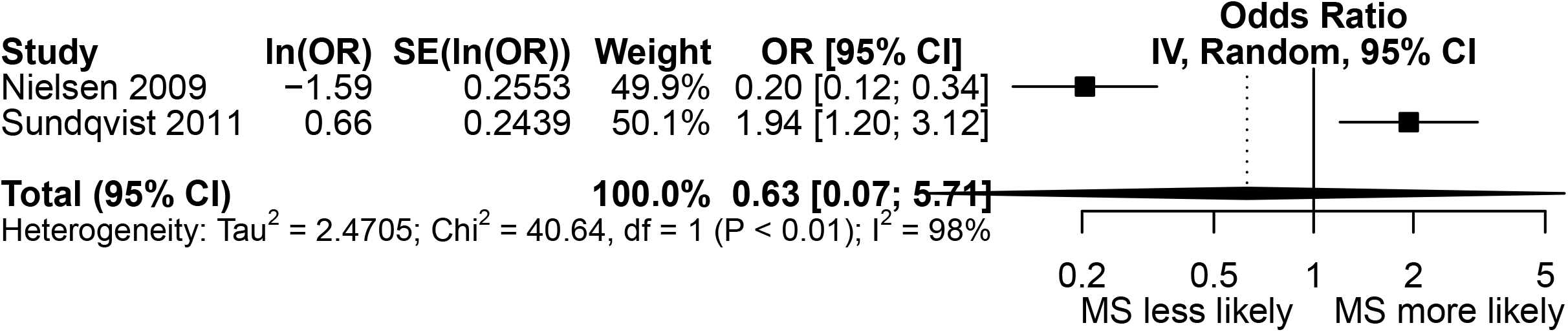

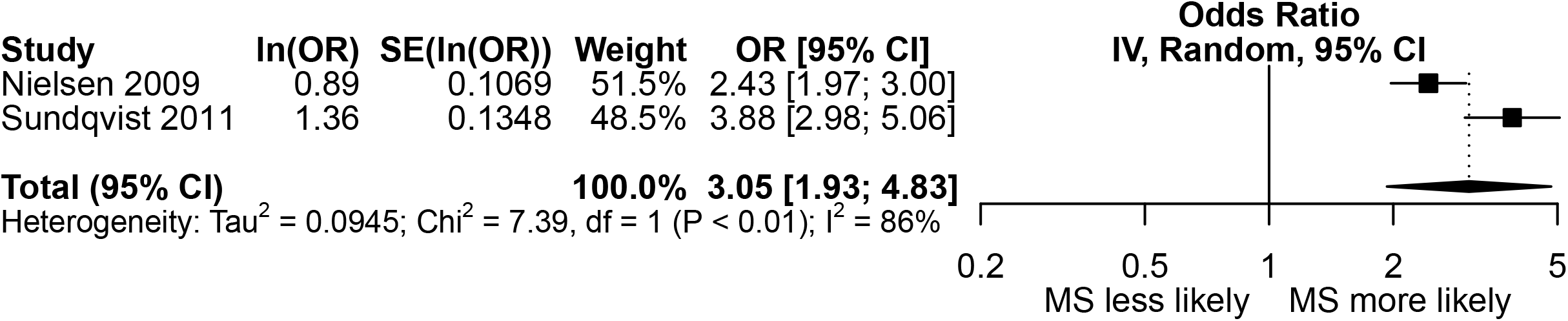

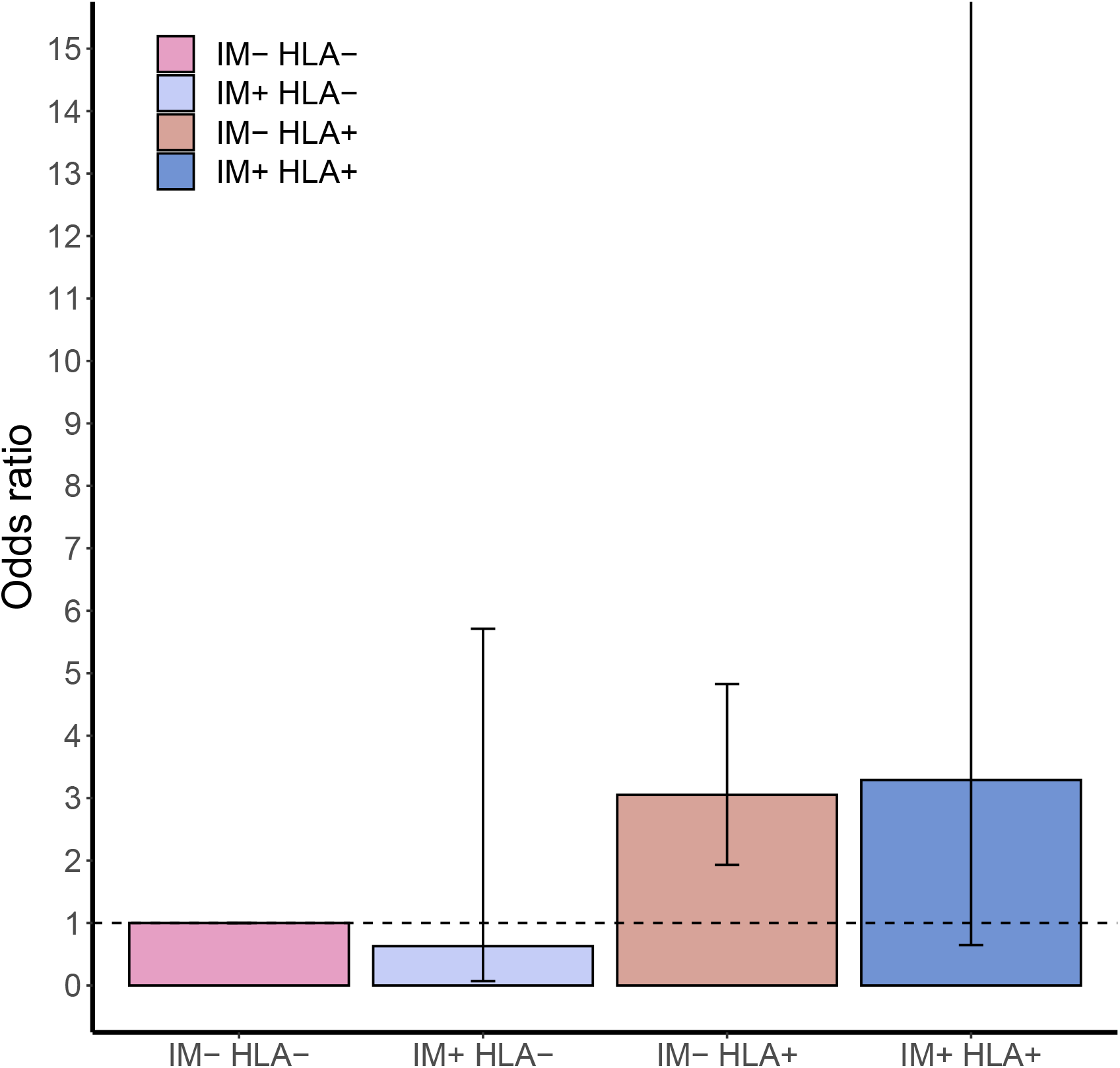

